# Genetic and transcriptomic determinants of disseminated coccidioidomycosis identify a founder variant in *NLRX1* and ancestry-specific rare variants in immune response genes

**DOI:** 10.64898/2026.06.26.26356412

**Authors:** Samantha L. Jensen, Sarah J. Spendlove, Alexis V. Stephens, Zhenjie Jin, Varada Abhyankar, Kangcheng Hou, Rachel Mester, Traci Toy, Eleazar Eskin, George R. Thompson, Royce H. Johnson, Arash Heidari, Rasha Kuran, Paul Krogstad, Bogdan Pasaniuc, Harold Pimentel, Manish J. Butte, Valerie A. Arboleda

## Abstract

Coccidioidomycosis, also known as Valley Fever, is a fungal disease endemic to the Americas that kills hundreds annually, yet the host factors that lead to increased risk of life-threatening dissemination of coccidioidomycosis remain poorly understood. We assembled the largest comprehensively sequenced coccidioidomycosis cohort to date, comprising 795 individuals with laboratory confirmed coccidioidomycosis and clinical disease severity phenotyping, many with paired whole blood genomic and transcriptomic data. Individuals with greater than 50% African genetic ancestry have increased risk for disseminated coccidioidomycosis (DCM) cases (OR=13.37, p=1.08×10^-18^), reflecting ancestry-associated differences in allele frequencies at immune loci. Transcriptomic profiling (n=267) revealed upregulation of interferon-inducible genes *IFI44* and *IFI44L*, the fungal recognition receptor *CLEC4D*, and pro-inflammatory protein *S100A12*, with sex-specific expression differences in immune cell composition. Gene-burden testing identified NOD-like receptor NLRX1 as the only gene carrying significantly more damaging rare variants than expected by chance (p=5.85×10⁻⁴). We identified a rare missense variant, *NLRX1* p.Arg252Trp (rs145644388), in five patients with DCM that represents a founder variant: all carriers share African local genetic ancestry and carry 0.6–1.1 centimorgans of identical-by-descent sequence, indicating origin from a common ancestor. In gnomAD, *NLRX1* p.Arg252Trp shows has higher allele frequency in African (AF=0.00615) compared to European (AF= 2.25x10^-5^) populations, directly linking this rare variant to population-level African genetic ancestry enrichment in DCM. *NLRX1* disruption impairs LC3-associated phagocytosis, an antifungal mechanism in macrophages. Together, these findings reveal both immune gene expression dysregulation and rare-variant architectures associated with African genetic ancestry underlying severe coccidioidomycosis and identify new targets for risk stratification and treatment.

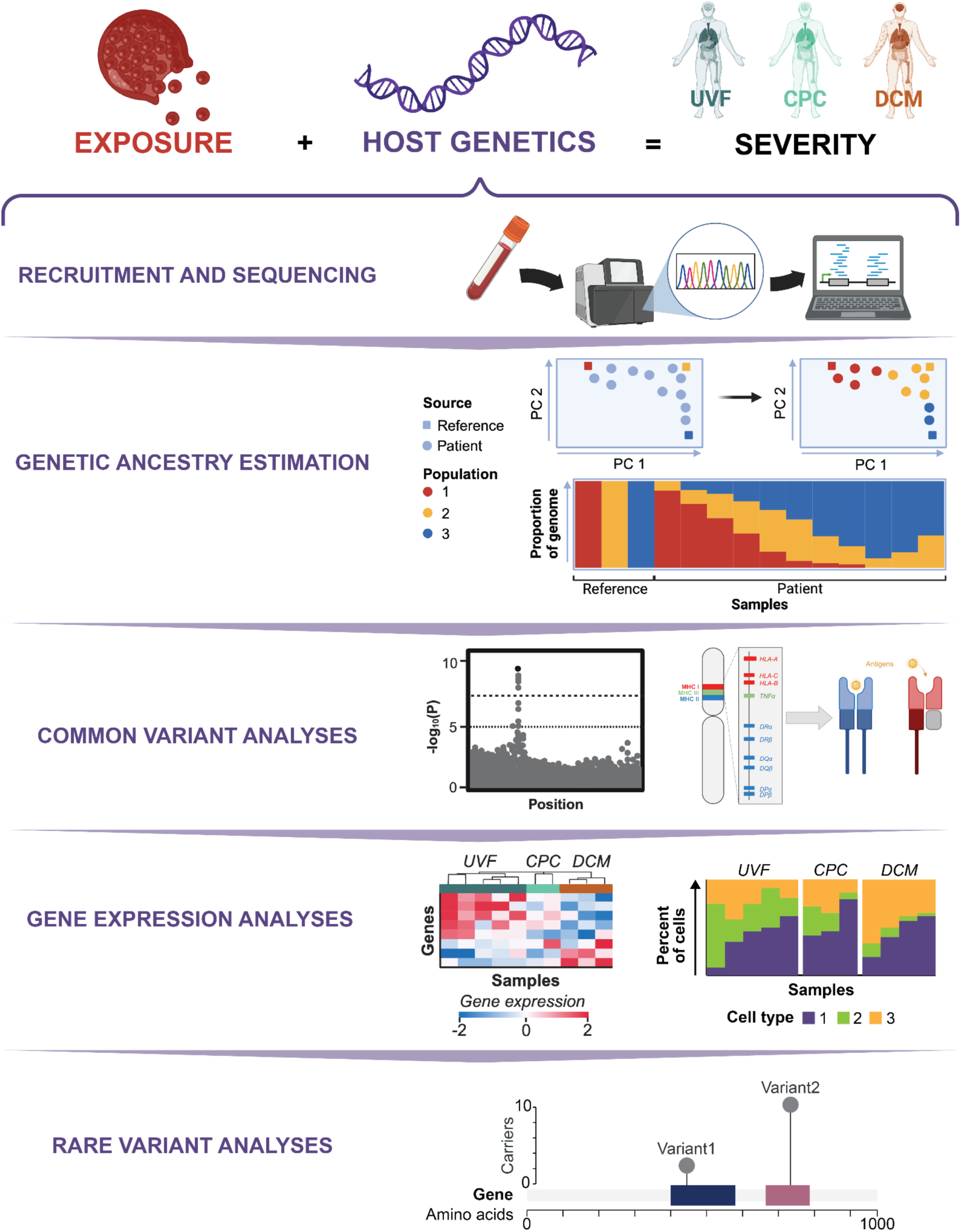

**Highlights:**

- Patients with disseminated coccidioidomycosis are significantly more likely to have African genetic ancestry.
- Whole-blood transcriptomics identifies upregulation of interferon-inducible genes *IFI44*, *IFI44L*, and fungal pattern-recognition receptor *CLEC4D* in disseminated disease.
- Rare missense variants in *NLRX1*, a mitochondrial NOD-like receptor involved in LC3-associated phagocytosis, are significantly enriched in disseminated cases by gene-burden testing.

**Context and Significance:** Valley fever (coccidioidomycosis) is a fungal infection endemic to the southwestern United States that causes life-threatening disseminated disease in a small fraction of those infected. The biological determinants underlying why some develop severe, disseminated infection remains poorly understood. Epidemiological studies have noted that individuals of African American or Filipino background face disproportionately higher risk for severe disease, but these studies relied on self-reported race, a social construct that is not biologically based. By assembling the largest genomically characterized Valley Fever cohort to date and using genome sequencing to directly quantify genetic ancestry, we show that genetic variation that is more common in populations with African ancestry is associated with risk for dissemination. We further identify disruption of interferon signaling and LC3-associated phagocytosis — a cellular mechanism by which macrophages contain fungal infections — as likely contributors to severe disease. These findings open new avenues for risk stratification and potential therapeutic targeting in this neglected fungal infection.

## INTRODUCTION

Coccidioidomycosis, also known as San Joaquin Valley Fever (or Valley Fever), is a fungal infection endemic to the southwestern United States, Central America, and South America. Primary infection occurs through inhalation or direct inoculation of arthroconidia, the fungal spores of *Coccidioides*. Since most infections are mild and self-resolving before laboratory diagnosis, the reported prevalence is underestimated. Disease severity is variable, ranging from unrecognized or mild respiratory symptoms — referred to here as uncomplicated valley fever (UVF) — to more complex pulmonary disease, termed complicated pulmonary coccidioidomycosis (CPC)^1–4^. In fewer than 1% of infections, *Coccidioides* spread outside of the lungs to bone, brain, skin, and other tissues, resulting in disseminated coccidioidomycosis (DCM)^1,2,5–7^. DCM is associated with significant morbidity and mortality and requires intensive and often lifelong treatment with antifungal medications^2,8,9^. Identifying biological risk factors associated with the more severe forms of coccidioidomycosis — CPC and DCM — is critical for improving both prevention strategies and therapeutic interventions^10–12^.

Interindividual differences in the host immune response have been shaped by co-evolution of human populations and endemic infectious pathogens. Over evolutionary time, alleles that confer protection against severe disease can increase in frequency due to the selective pressure of the pathogen, and despite the deleterious effects of these variants. A classic example is the sickle cell variant, p.Glu7Val in β-hemoglobin (HBB), where heterozygosity provides protection against malaria parasitemia^13,14^ and is present at allele frequencies exceeding 9% in parts of Africa^15^ but homozygosity causes sickle cell disease. Another source of ancestry-specific variation is due to polygenic adaptation, in which many allele frequencies in a population change in response to a selective pressure. An increased proportion of European genetic ancestry is associated with strong type I interferon pathway activity in response to influenza infection, and this effect was driven by genetic ancestry associated differences in allele frequency at cis-regulatory variants^16^. Ancestry-specific alleles associated with pathogen sensing in dendritic cells are linked to increased host inflammatory responses^17^. The cell-type and pathogen-specificity of these effects have been identified for tuberculosis, where Peruvians with higher proportions of Native American genetic ancestry have a threefold increased risk of tuberculosis progression, after controlling for socioeconomic factors^18^. Overall, genetic variation, shaped in part by a population’s exposure to pathogens, can modulate immune cell signaling and contribute to differences in host susceptibility and response to infection.

Epidemiological risk factors for DCM include male sex, increased age, occupational exposure to dust in endemic regions^19–25^, and self-identified race or ethnicity (SIRE) groups^26–28^, with African American, Latino, and Filipino SIRE consistently overrepresented among the most severe cases. However, SIRE is a socio-political construct that is not an accurate surrogate for genetically defined ancestry^29,30^ and its use as a biological variable conflates social determinants of health with biological factors.

Previous studies identified associations between blood types and DCM risk in Hispanic populations^31^ and between human leukocyte antigen (HLA) alleles and DCM severity^32^. However, these studies were both underpowered and relied on SIRE, limiting their biological interpretability since HLA is strongly correlated with genetic ancestry^33^. Using genomic information, our study directly addresses these limitations by using genome sequencing data to quantify continental ancestry and immune loci associated with dissemination risk.

Both common and rare variants could influence risk of DCM. To date genome-wide association studies (GWAS) for coccidioidomycosis have not found any genome-wide significant loci^34^. Rare variant analyses have been more successful, identifying coding variation in pattern-recognition receptor *CLEC7A*, in cytokine receptors *IFNGR1*, *IL12RB1*, and *IL12RB2*, and in immune signaling genes, like *STAT1* and *STAT3*, that are associated with DCM^2,35–40^.

In this study, we assembled the largest coccidioidomycosis patient cohort to date, comprising 795 individuals with confirmed infection and detailed phenotyping across a wide spectrum of disease severity. We found that individuals with a high proportion of African genetic ancestry are significantly overrepresented among DCM cases. Differential expression analysis of RNA-seq data from a subset of participants (n=267) revealed marked disruption of immune-related pathways in individuals with CPC and DCM. While evaluation of common variant contributions was underpowered, we identified 1,327 rare variants of interest in immune response genes. One gene, NOD-like receptor *NLRX1*, was significantly associated with disseminated disease in a gene-based test and harbored two rare missense variants predicted to disrupt protein function that were only carried by individuals with severe disease (5 with DCM, 1 with CPC). Collectively, our findings identify genetic loci associated with coccidioidomycosis severity and offer new insight into biological mechanisms that may improve prognostic evaluation in chronic and disseminated disease.

## RESULTS

### Genomic biobank of patients with laboratory diagnosis of coccidioidomycosis

A challenge in coccidioidomycosis research is the development of a well-phenotyped cohort drawn from endemic regions. Up to 90% of exposed individuals remain asymptomatic or experience only mild, self-limiting symptoms, resulting in substantial underrepresentation of mild disease in previous studies. To overcome this, we assembled a cohort of patients with laboratory-confirmed coccidioidomycosis, recruited from three endemic California sites: the University of California, Davis; the University of California, Los Angeles; and the Valley Fever Institute at Kern Medical Center in Bakersfield (**Fig. 1A**). This is the largest and most comprehensively sequenced cohort with laboratory-diagnosed coccidioidomycosis, clinical disease severity phenotyping, and paired genomic and whole blood transcriptomic sequencing.

**Figure 1:**
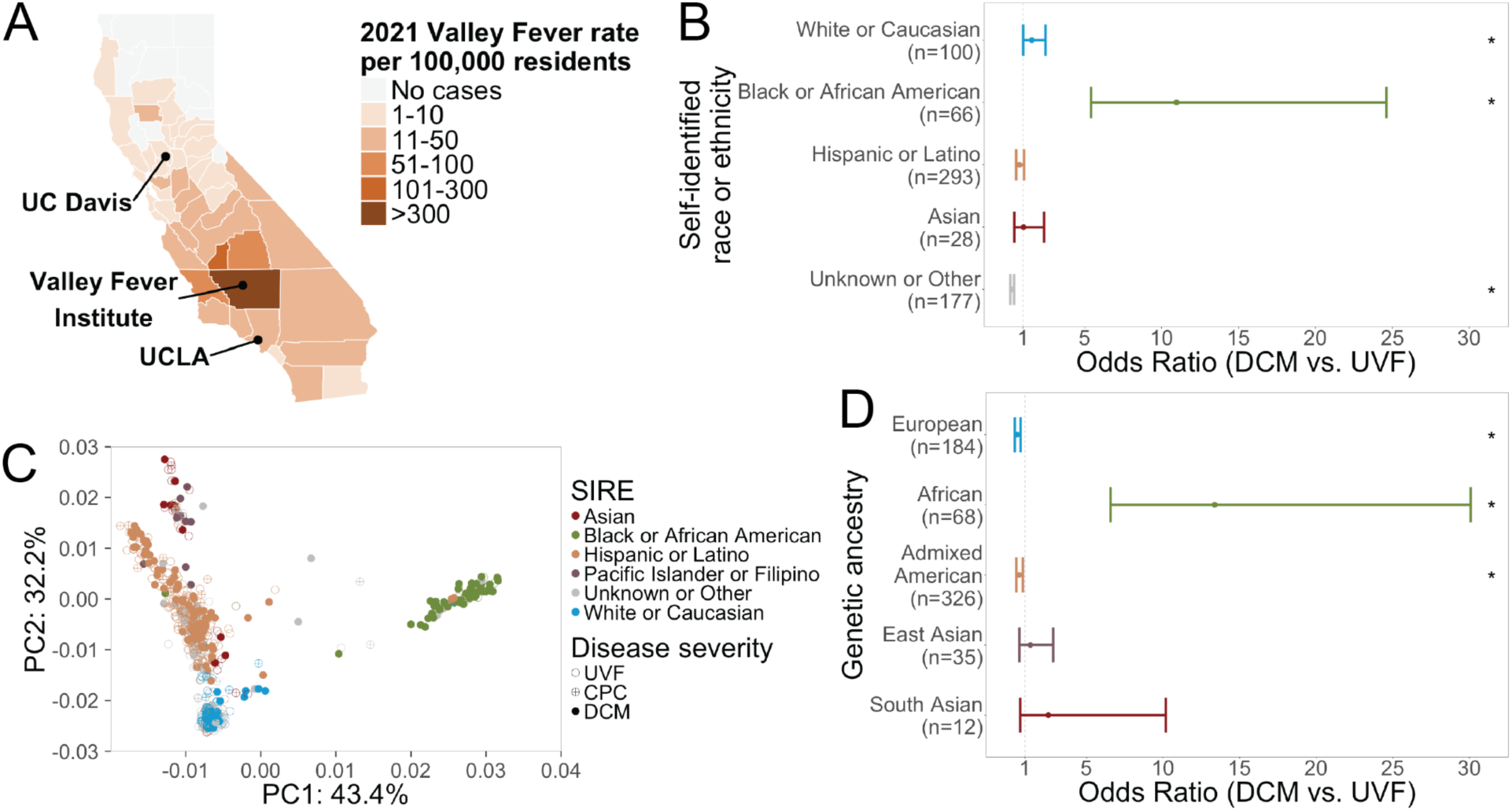
SIRE and genetic ancestry are distinct but significantly associated with risk for disseminated coccidioidomycosis. (A) Coccidioidomycosis patients were recruited from three endemic California sites: the University of California Davis, UCLA Health, and the Valley Fever Institute at Kern Medical Center in Kern County. Coccidioidomycosis incidence for all California counties is shown. Source: California Department of Public Health. (B) Odds ratios (ORs) for DCM versus UVF by self-identified race/ethnicity (SIRE), with each group compared to all other SIRE groups combined. Participants identifying as "Black or African American" had significantly higher odds of DCM (OR=10.97, p=6.02×10⁻¹⁶, Fisher’s exact test). No participants in the "Pacific Islander or Filipino" category had UVF (n=11, p=6.81×10⁻⁵), so the odds ratio was undefined. Significant comparisons (p <0.05) are marked with an asterisk. (C) Principal components analysis (PCA) conducted with common variants (AF >0.05) from DNA-sequenced coccidioidomycosis samples (n=733) and reference samples (n=3,356) from the 1000 Genomes Project, HGDP, and SGDP. Filled and open circles represent DCM and non-DCM participants, respectively. Most participants with "Black or African American" SIRE (green) clustered within the African genetic ancestry region (PC1 > 0.01). Among participants with primarily African genetic ancestry (n=80), 73% had DCM — significantly higher than the 25% DCM rate among all other patients (OR=7.59 [95% CI: 4.42–13.45], p=3.88×10⁻¹⁶, Fisher’s exact test). (D) Odds ratios for DCM by primary genetic ancestry group, each compared to all other ancestry groups combined. Patients with European (OR=0.48 [95% CI: 0.32–0.72], p=1.73×10⁻⁴) or admixed American (OR=0.61 [95% CI: 0.43–0.86], p=3.50×10⁻³) genetic ancestry were significantly less likely to have DCM. Patients with African genetic ancestry had significantly higher odds of DCM (OR=13.37 [95% CI: 6.57–30.07], p=1.08×10⁻¹⁸).

In total, we recruited 795 individuals: 413 with UVF, 121 with CPC, and 261 with DCM. The male (n=510) to female (n=285) ratio of 1.8 is consistent with studies showing that males are 1.8-fold more likely to have symptomatic coccidioidomycosis^25^ (**Table 1**, Sex).

**Table 1:** Demographics of California coccidioidomycosis cohort. This table contains a summary of demographic and clinical characteristics of the 795 participants included in analyses, stratified by disease severity (UVF, CPC, DCM). The male-to-female ratio, self-identified race/ethnicity distribution, and genetic ancestry proportions in our cohort are consistent with those reported in prior epidemiological studies of coccidioidomycosis. For each trait, the number of individuals who had non-missing values is marked (N). For categorical variables, the Overall, UVF, CPC, and DCM columns contain the number of samples with a given trait and the percent of the total samples that entails in parentheses. For continuous variables these columns contain the median value and the first and third quartile values in parentheses. The *p*-value for each comparison is calculated with a Pearson’s Chi-squared test for categorical variables, with a Kruskal-Wallis rank sum test for continuous variables like age and proportion of African genetic ancestry, and a Fisher’s exact test with simulated p-value based on 2,000 replicates for categorical count data like SIRE and major genetic ancestry.

| Trait | Value | Overall | UVF<br>n=413<br>(52%) | CPC<br>n=121<br>(15%) | DCM<br>n=261<br>(33%) | p-value |
| --- | --- | --- | --- | --- | --- | --- |
| Sex (N=795) | Female | 285<br>(36%) | 184<br>(23%) | 41<br>(5.2%) | 60<br>(7.5%) | <0.001 |
|  | Male | 510<br>(64%) | 229<br>(29%) | 80<br>(10%) | 201<br>(25%) |  |
| Age (N=790) |  | 57<br>(41, 67) | 58<br>(51, 69) | 56<br>(40, 69) | 55<br>(41, 63) | 0.088 |
| Self-identified race or ethnicity (N=795) | Asian | 33<br>(4.2%) | 17<br>(2.1%) | 5<br>(0.6%) | 11<br>(1.4%) | <0.001 |
|  | Black or African American | 76<br>(9.6%) | 10<br>(1.3%) | 10<br>(1.3%) | 56<br>(7.0%) |  |
|  | Hispanic or Latino | 350<br>(44%) | 190<br>(24%) | 57<br>(7.2%) | 103<br>(13%) |  |
|  | Native American | 1<br>(0.1%) | 0<br>(0%) | 1<br>(0.1%) | 0<br>(0%) |  |
|  | Pacific Islander or Filipino | 11<br>(1.4%) | 0<br>(0%) | 1<br>(0.1%) | 10<br>(1.3%) |  |
|  | Unknown or Other | 203<br>(26%) | 144<br>(18%) | 26<br>(3.3%) | 33<br>(4.2%) |  |
|  | White or Caucasian | 121<br>(15%) | 52<br>(6.5%) | 21<br>(2.6%) | 48<br>(6.0%) |  |
| Major genetic ancestry (N=733) | Admixed American | 377<br>(51%) | 226<br>(31%) | 51<br>(7.0%) | 100<br>(14%) | <0.001 |
|  | African | 80<br>(11%) | 10<br>(1.4%) | 12<br>(1.6%) | 58<br>(7.9%) |  |
|  | East Asian | 41<br>(5.6%) | 20<br>(2.7%) | 6<br>(0.8%) | 15<br>(2.0%) |  |
|  | European | 222<br>(30%) | 138<br>(19%) | 38<br>(5.2%) | 46<br>(6.3%) |  |
|  | South Asian | 13<br>(1.8%) | 5<br>(0.7%) | 1<br>(0.1%) | 7<br>(1.0%) |  |
| Proportion African ancestry (N=733) |  | 0.04<br>(0.01,0.07) | 0.03<br>(0.00, 0.06) | 0.03<br>(0.01, 0.07) | 0.05<br>(0.01, 0.65) | <0.001 |

### SIRE-based associations with DCM are reproducible but limited by missing data and biological imprecision

Multiple independent studies have concluded that African American, Latino, or Filipino individuals are at increased risk for severe coccidioidomycosis infections and dissemination^21,41–44^. We harmonized six coccidioidomycosis cohort studies (1944–2008, total n=1,529)^21,41–45^ and a meta-analysis showed that non-white individuals were more likely to have disseminated disease (OR=11.47 [95% CI: 7.77–17.15], *p*=1.59×10⁻⁴⁰, Fisher’s Exact Test for Count Data, **Fig. S1A**). Those who self-identified as ‘African American or Black’ had over five-fold higher odds to have DCM than individuals of all other SIRE groups (OR=5.33 [95% CI: 3.55-7.94], p=1.26x10^-15^). Individuals who identified as Filipino had over nine times the odds of DCM compared to all other SIRE groups (OR=9.14 [95% CI: 5.34-15.55], *p*=1.96x10^-15^).

In our new cohort of 795 individuals, we first wanted to assess how well the SIRE demographic data matched our meta-analysis of previously published cohorts (**Fig. S1A**). In our cohort, participants who self-reported as “Black or African American” were overrepresented (**Table 1**, Self-identified race or ethnicity), comprising 9.6% of our cohort compared with the reported 5.4% in California for the 2020 census^46^. ‘Black or African American’ SIRE (OR = 10.97 [95% CI: 5.40-24.62], *p*=6.02x10^-16^, **Fig. 1B**) and ‘Pacific Islander or Filipino’ SIRE (*p*=6.81x10^-5^) were each significantly associated with DCM. While this data revealed some interesting trends, 25.7% (204/795) of our cohort did not have an assigned SIRE, and this missing data highlights the limitations of relying on SIRE.

### Genetic ancestry is associated with coccidioidomycosis dissemination risk

We generated whole genome or exome sequencing and used this to infer each participant’s genetic ancestry, which is an individual’s genomic similarity to known global reference populations^47–49^. Genetic data allowed us to distinguish genetically mediated disease risk from environmental risk factors that co-vary with SIRE. Principal components analysis (PCA) with references from known populations (**Fig. S1B**) showed that individuals with African genetic ancestry made up 11% (80/733) of our population but 26% (58/226) of DCM patients (**Fig. 1C** and **Table 1**, Major genetic ancestry). Within each of the SIRE categories, SIRE labels are not perfectly correlated with genetic ancestry based on PCA: For example, several individuals who identified as “Latino or Hispanic” have primarily African genetic ancestry (**Fig. S2, Fig. 1C**).

Individuals with primarily European genetic ancestry had decreased odds of dissemination compared with all other major continental genetic ancestry groups (**Fig. 1D**, OR=0.48 [95% CI: 0.32-0.72], *p*=1.73x10^-4^, Fisher’s Exact Test). Individuals with African genetic ancestry were more likely to have DCM (OR=13.37 [95% CI: 6.57-30.07], *p*=1.08x10^-18^). We also find that individuals with high proportions of admixed American genetic ancestry were less likely to have disseminated disease (OR=0.61 [95% CI: 0.43-0.86], *p*=3.50x10^-3^). Our data highlight that nuanced assessments using genetic ancestry similarity can improve power to detect associations that may reflect true biological signals.

### Continuous genetic ancestry estimation reveals African ancestry enrichment in Disseminated Coccidioidomycosis

To move beyond discrete ancestry categories, we applied an unsupervised ADMIXTURE model^50^ to generate continuous estimates of continental genetic ancestry for each individual (**Fig. 2A**). An increased proportion of African genetic ancestry is associated with increased risk of DCM (**Table 1**, Estimated proportion African ancestry). Individuals with more than 50% African genetic ancestry represent 25.2% (57/226) of the DCM patients compared with just 11.1% (12/108) of CPC patients and 2.3% (9/399) of the UVF population (**Fig. 2B**). In other words, the proportion of our cohort with more than 50% of their genome estimated to be of African genetic ancestry is 10 times greater in DCM than in UVF (*p*=3.35x10^-13^, Welch Two Sample *t*-test). DCM patients had 19% more variants associated with African ancestry than those with UVF (**Figure 2C**, *p*=6.81x10^-14^, Welch two-sample *t*-test).

**Figure 2:**
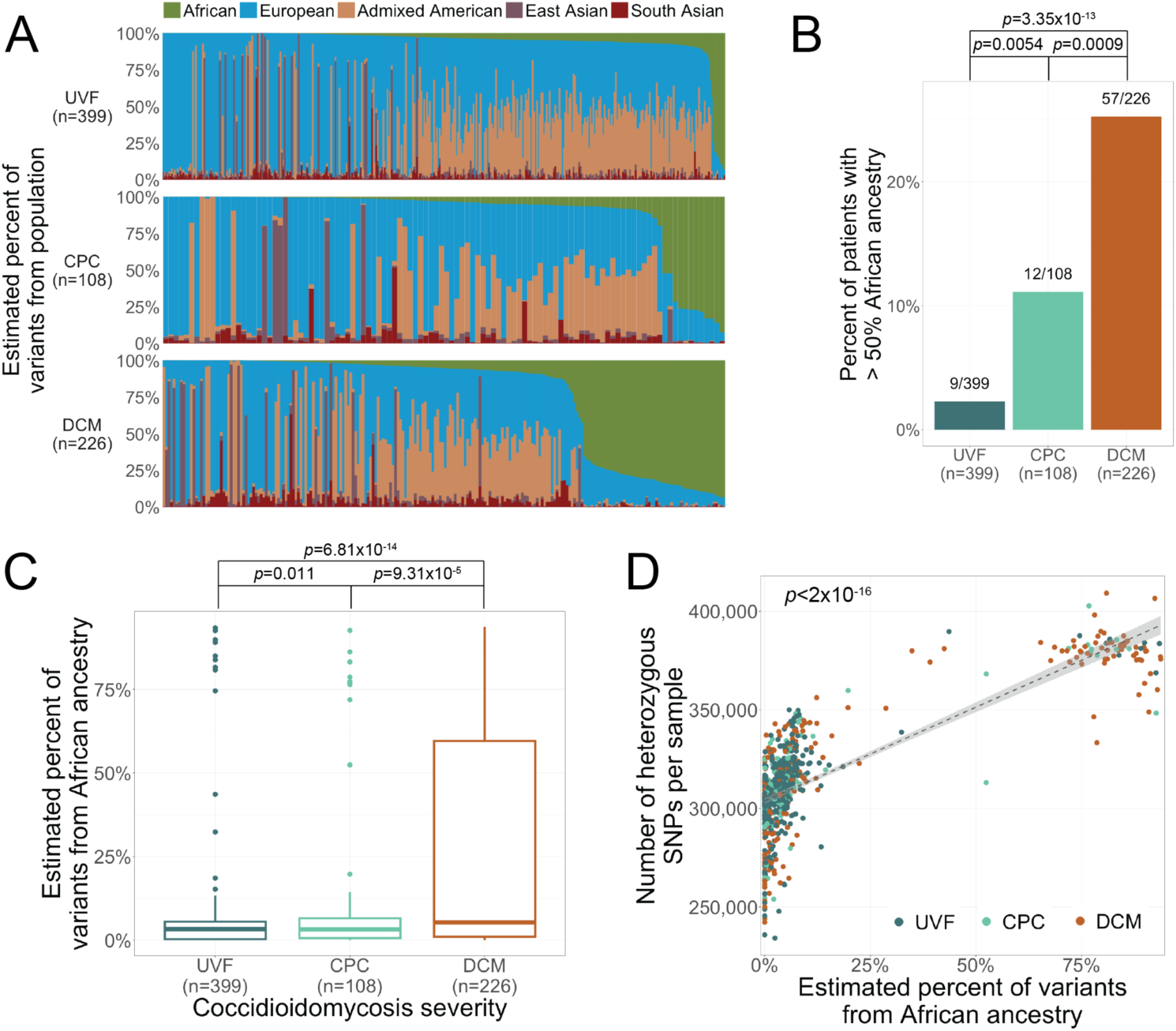
Continuous estimation of genetic ancestry reveals dose-dependent enrichment of African ancestry in disseminated coccidioidomycosis. (A) Global genetic ancestry proportions estimated for each individual using an unsupervised ADMIXTURE model (k=5) with reference populations. Each bar represents one individual; bar segments indicate the estimated proportion of each continental ancestry group. Individuals are ordered left to right by increasing African genetic ancestry (green). Individuals with DCM (bottom row) are enriched for higher African genetic ancestry proportions relative to those with CPC and UVF. (B) Individuals with more than 50% African genetic ancestry represent 25.2% (57/226) of DCM patients, compared with 11.1% (12/108) of CPC patients (p=8.8×10⁻⁴, Welch two-sample t-test) and 2.3% (9/399) of UVF patients (*p*=3.35×10⁻¹³). (C) Mean estimated African genetic ancestry is significantly higher in DCM patients (24%) than in those with CPC (11%, *p*=9.31×10⁻⁵) or UVF (5%, *p*=6.81×10⁻¹⁴). All comparisons by Welch two-sample t-test. (D) DCM patients have significantly more heterozygous SNPs than those with UVF (mean 325,769 vs. 309,153, *p*=1.28×10⁻¹¹, linear regression) or CPC (mean 315,675, *p*=0.003). For each additional percentage point of African genetic ancestry, patients carry an estimated 952 additional heterozygous SNPs (*p*<2×10⁻¹⁶, linear regression), consistent with greater genetic diversity in African populations.

For each additional percentage point of estimated African global genetic ancestry, participants had an average of 952 more heterozygous SNPs (**Fig. 2D**, *p*<2x10^-16^, linear regression), which is consistent with known increased genetic diversity in African populations. Conversely, the estimated percent of European genetic ancestry was negatively correlated with both risk of dissemination (**Fig. S3A-C**, 19.36% more European genetic ancestry in UVF than DCM [95% CI:14.63%-24.10%], *p*=7.82x10^-15^, Welch two-sample t-test) and genetic diversity (**Fig. S3D**, β=-127 SNPs, *p*=4.5x10^-4^, linear regression).

### Genome-wide association study is underpowered to detect loci contributing to DCM risk

A prior GWAS, which included a subset of our cohort, identified no genetic loci significantly associated with coccidioidomycosis severity^34^, however we reasoned that the addition of 267 new individuals might improve power sufficiently to detect large-effect common variants, if such variants exist. We performed GWAS using logistic regression correcting for age, sex, and the first ten principal components (see **Methods**, **Figure S4A**). No loci reached genome-wide significance (p < 5×10⁻⁸). We identified five nominally significant loci containing seven variants (p < 1×10⁻⁵, **Table S1**, **Figure S4B**), all noncoding and located in intronic (6/7) or intergenic (1/7) regions. The number of nominally significant hits is consistent with chance expectation under the null (**Figure S4C**), and these results should be interpreted accordingly.

One nominally significant variant, rs10845753, an intergenic SNP downstream of *DPPA3*, is most common in Asian and European genetic ancestry. It alters 19 different regulatory motifs, including transcription factor binding motifs for histone deacetylase 2 (*HDAC2*), a transcriptional repressor involved in major histocompatibility complex (MHC) II repression in COVID^51^, and interferon regulatory factor 9 (*IRF9*), which complexes with *STAT1* and *STAT2* to form the IFN-stimulated gene factor 3 (ISGF3) complex in response to signaling from the IFN-α receptor^52,53^. While the convergence of this signal with the interferon pathway dysregulation is biologically interesting, this variant does not reach genome-wide significance and its association with DCM requires replication before any functional inference is warranted.

### HLA type is associated with DCM but strongly correlated with genetic ancestry

HLA allele frequencies differ substantially across ancestral groups, so unadjusted HLA associations with DCM are expected to partly reflect ancestry stratification. To test whether any MHC class II allele — the genomic region encoding HLA proteins — was independently associated with disease severity, we called HLA types in 626 samples. In an initial unadjusted screen (Fisher’s Exact Test, p < 4.0×10⁻⁴), two alleles were more common in DCM patients than UVF (DPA1*03:03 and DRB1*15:03) and one allele (DPA1*01:03) was less common (**Figure S5A**).

DRB1*15:03 was previously found to be more common in African American patients with severe coccidioidomycosis than in outside controls in Kern County, California^32^. Consistent with this, the DRB1*15:03 allele frequency in DCM patients was ten-fold higher than in participants with UVF (p=1.29×10⁻⁷, OR=11.12, 95% CI: 3.74–44.72) and was elevated in CPC patients relative to UVF as well (p=0.024, OR=4.77, 95% CI: 1.02–24.31).

DPA1*01:03 showed an apparent protective pattern: while it accounted for ∼80% of DPA1 alleles in UVF and CPC patients, it comprised only 64.5% of DCM patients’ alleles (UVF vs. DCM p=6.83×10^⁻⁹^; CPC vs. DCM p=1.57×10^⁻³^, Figure S5B). However, DPA1*01:03 is the dominant allele in admixed American and European populations (AF >75%) but comprises less than 50% of alleles in African populations (**Figure S5C**)^54^. The apparent protective effect of DPA1*01:03 therefore likely reflects its lower frequency in individuals with African genetic ancestry rather than an independent HLA effect. Indeed, after correcting for age, sex, sequencing batch, and genetic ancestry in logistic regression, no MHC class II allele was significantly associated with disease severity (FDR <0.05, **Table S2**), confirming that the unadjusted HLA signals are driven by ancestry stratification and not an independent HLA effect.

### Whole-blood transcriptomics reveals dysregulation of interferon and antifungal immune pathways in DCM

To further explore the molecular underpinnings of DCM risk, we performed whole blood transcriptomic analysis on a subset of our cohort (n=267) using blood samples obtained at the enrollment visit. We performed differential gene expression analysis between DCM (n=122) and UVF (n=100), correcting for age, sex, and sequencing batch, and identified 12 genes that were significantly upregulated in DCM patients (**Fig. 3A**, **Table S3**). Seven of the twelve significantly differentially expressed genes were immune related genes^55–57^ and we performed functional enrichment analysis on the nine protein-coding differentially expressed genes using the STRING database and identified multiple enriched annotations (FDR<0.05, **Table S4**). Immune annotations, including ‘Immune response’ and ‘Defense response to fungus’, were significantly enriched (FDR<0.023, **Fig. 3B**).

**Figure 3:**
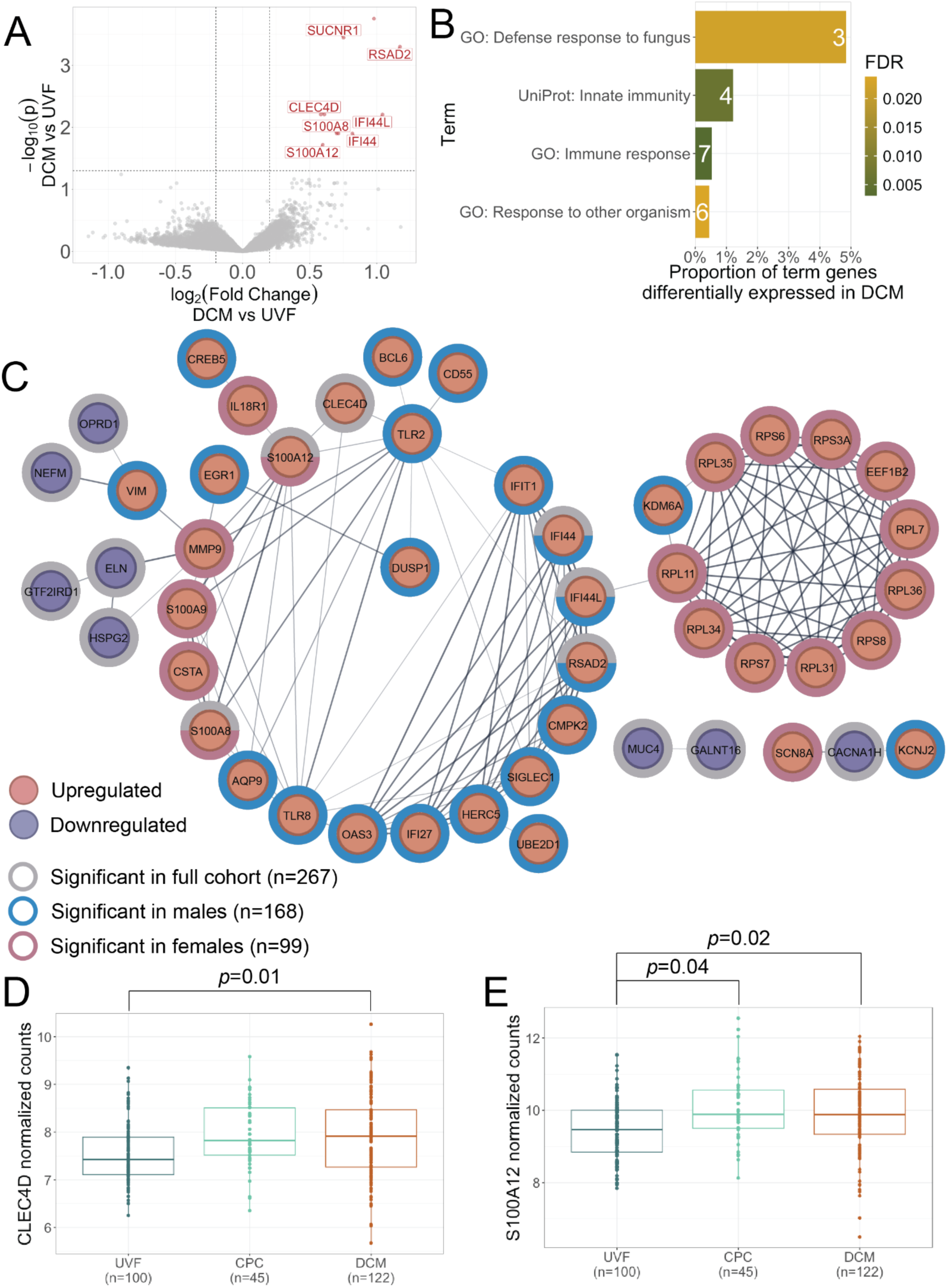
Transcriptomic analysis identifies dysregulation of interferon signaling and antifungal immune pathways in DCM. (A) Volcano plot of whole blood differential gene expression between participants with DCM (n=122) and those with UVF (n=100). Twelve genes were differentially expressed (DE) in DCM compared to UVF (adjusted *p*<0.05) and showed increased expression (red dots) in DCM patients. The labeled genes have established immune function. (B) Using the STRING database, we annotated the nine protein-coding DE genes and tested for functional enrichment. This method predicted more protein-protein interactions (PPIs) than would be expected by chance (*p*=2.9x10^-6^). Six genes had at least one PPI with another DE gene based on annotations in databases or literature, co-expression, or protein homology. Three Gene Ontology (GO) biological process terms and one UniProt annotated keyword term, all associated with host immunity, were significantly enriched (*p* < 0.02). (C) Protein-protein interaction network of all genes differentially expressed in any pairwise comparison. Edge thickness represents strength of PPI evidence. Node color indicates direction of differential expression; ring color indicates the comparison in which significance was reached: the full cohort (grey ring), male patients only (blue ring), or female patients only (pink ring). Interferon signaling genes predominate among male-specific findings (left); genes encoding ribosomal proteins are enriched among female-specific findings (right). (D) *CLEC4D*, a paralog of the known coccidioidomycosis risk gene *CLEC7A*, was significantly upregulated in DCM patients relative to UVF (*p*=0.01). (E) *S100A12* was the only gene significantly upregulated in both DCM and CPC patients relative to UVF (*p*=0.02 and *p*=0.04, respectively).

Six of the differentially expressed genes are predicted to interact with each other (**Fig. 3C**), spanning two aspects of host immunity: type I interferon signaling and fungal immune defense. The type I interferon (IFN) signaling genes include *interferon-induced protein 44* (*IFI44*), *interferon-induced protein 44-like* (*IFI44L*), and *radical S-adenosyl methionine domain containing 2* (*RSAD2*). *IFI44* encodes an IFN-α-inducible protein implicated in susceptibility to COVID-19*, Staphylococcus aureus,* hepatitis C, and other infectious pathogens^58–62^. *IFI44L*, a nearby paralog, has been implicated in autoimmune disorders including systemic lupus erythematosus and rheumatoid arthritis, and modulates the strength of the immune response to tuberculosis and childhood respiratory tract infections^62–66^. The remaining three interacting genes — *C-type lectin domain family 4 member D* (*CLEC4D*), *S100 calcium binding protein A8* (*S100A8*), and *S100 calcium binding protein A12* (*S100A12*) — are functionally annotated with the gene ontology (GO) term ’immune defense response to fungus’. *CLEC4D*, a paralog of the known coccidioidomycosis risk gene *CLEC7A*^35,67–69^, was significantly upregulated in DCM patients (*p*=0.01, **Fig. 3D**) and encodes pattern recognition receptor Dectin-3, which complexes with Dectin-2 to recognize α-mannans in *C. albicans* and initiates T-cell priming toward T-helper 1 and T-helper 17 subtypes^70,71^.

We next compared gene expression profiles between CPC (n=45) and UVF (n=100) patients and identified 19 significantly differentially expressed genes, most of which were downregulated relative to UVF (**Fig. S6A**). Only one gene, *S100A12*, was upregulated in both DCM and CPC relative to UVF (*p*=0.02 and *p*=0.04, respectively, **Fig. 3E**). *S100A12* encodes a calcium-binding pro-inflammatory protein implicated in lung inflammation and upregulated in patients with allergic asthma^72,73^. Increased *S100A12* expression has also been associated with greater severity of influenza, COVID-19, and community-acquired pneumonia^74,75^.

### Interferon signaling dysregulation in severe coccidioidomycosis is most pronounced in male patients

Because DCM is more common in male patients^25^, we also assessed differential expression separately in males and females (**Figure 3C** and **Table S3**). In males (n=168), 13 genes were upregulated in DCM relative to UVF (**Figure S6B**), 22 in CPC relative to UVF (**Figure S6C**), and 3 in DCM relative to CPC (**Figure S6D**). In males, three genes are upregulated in DCM relative to both UVF or CPC, *IFI44L*, *RSAD2*, and *sialic acid binding immunoglobulin like lectin 1* (*SIGLEC1*) (**Figure S7A**). Like *IFI44L* and *RSAD2*, *SIGLEC1* is involved in interferon signaling. Elevated *SIGLEC1* expression is associated with reduced IFN-ɣ signaling by T-cells and has been implicated in RSV, COVID-19, and tuberculosis pathogenicity^76–79^. Several other differentially expressed genes in males with DCM or CPC compared with UVF are involved in Type I or II interferon signaling including *IFN-α-inducible protein 27* (*IFI27*) and *IFN-induced protein with tetratricopeptide repeats 1* (*IFIT1*)^80–86^. Additionally, *high mobility group box 2* (*HMGB2*), *2’-5’-oligoadenylate synthetase 3* (*OAS3*), *toll like receptor 2* (*TLR2*), and *toll like receptor 8* (*TLR8*) positively regulate IFN-ɣ production (**Figure S7B**).

In females (n=99), 41 genes were differentially expressed in DCM relative to UVF: 9 downregulated and 32 upregulated (**Figure S6E**). An additional 3 genes, including immune-related gene *SEMA5A*, are upregulated in DCM relative to CPC (**Figure S6F**). Among genes upregulated in DCM relative to UVF are some related to IFN-ɣ signaling, like *interleukin 18 receptor 1* (*IL18R1*). Although female patients had the only significantly downregulated genes in DCM relative to UVF, only one gene - *T-cell receptor alpha joining 29* (*TRAJ29*) - was protein coding. Unexpectedly, almost a third (10/32) of the upregulated genes in female patients encode ribosomal proteins (**Figure S7B**), which may not have a clear functional significance or represent uncorrected technical artifacts. Across both the full cohort and sex-stratified models, differentially expressed genes converge on type I and II interferon signaling as a central axis of dysregulation in severe coccidioidomycosis, with the strongest signal observed in male patients (**Figure S8**).

### Immune cell deconvolution reveals B-cell depletion in DCM independent of sex

To explore whether the transcriptomic differences between groups reflected differences in immune cell composition, we used CIBERSORTx to deconvolve immune cell type proportions from our RNA-seq data^87^ (**Figure S9A**). Although the average differences were small, DCM patients had significantly fewer B cells (*p*=0.006, 95% CI: 0.0029–0.017, Welch two-sample t-test, **Figure S9B**) and T cells (*p*=0.025, 95% CI: 0.0031–0.0457, **Figure S9C**) than individuals with UVF.

Because sex hormones have known effects on immune cell proportions^88,89^ and male patients are overrepresented among those with CPC and DCM in our cohort, we tested whether the observed cell-type differences were confounded by sex (**Figure S9D**). Male patients had 3% fewer T cells than female patients (*p*=0.0015, Welch two-sample t-test). After correcting for sex, age, and RNA-seq batch, the association between disease severity and T-cell proportion was no longer significant (*p*=0.16, linear regression) and the T-cell depletion in DCM appears to at least partially reflect the greater proportion of male patients among severe cases rather than an independent effect of disease severity.

In contrast, there was no significant effect of sex on B-cell proportion (*p*=0.25, Welch two sample t-test), and DCM patients retained a significantly lower proportion of B cells after correcting for sex, age, and RNA-seq batch (*p*=0.023, β=−0.0089, linear regression). This is biologically notable: while T cells are critical for initial protection against *Coccidioides*, B cells contribute importantly to sustained immunity following exposure^90–93^.

### Rare variants in known coccidioidomycosis risk genes are enriched in patients with DCM

Rare genetic variation is a major driver of host immunodeficiency and response, which can lead to severe coccidioidomycosis outcomes. Rare coding variants in immune genes — including pattern-recognition receptor *CLEC7A*, cytokine receptors *IFNGR1*, *IL12RB1*, and *IL12RB2*, and immune signaling genes *STAT1* and *STAT3* — have previously been associated with severe coccidioidomycosis and immunodeficiency in disseminated cases^2,35,36,38–40,94^. We assessed DNA sequencing from 751 patients to identify genetic variants that could contribute to dissemination risk (**Figure S10A**). We identified 15 of the 36 variants previously associated with coccidioidomycosis in our cohort (**Figure S10B**, **Table S5**), and these variants were carried by 30.3% (228/751) of our cohort. The most observed variant were variants in *CLEC7A*, which we describe below, before turning to novel rare variant discovery across a broader set of immune response genes.

A *CLEC7A* nonsense variant, p.Tyr238Ter (rs16910526), is predicted to produce a truncated Dectin-1 protein. There are 3 homozygous and 66 heterozygous individuals in our cohort (**Figures S11A–B**). Although this variant is relatively common in the gnomAD population database (AF=0.073), its frequency in our cohort was significantly lower than expected (AF=0.047, Fisher’s Exact Test *p*=8.4×10⁻⁵, **Figure S11C**). However, within our cohort, rs16910526 was significantly enriched in those with DCM (AF=0.060, *p*=0.019) and CPC (AF=0.074, *p*=0.010). All individuals homozygous for this truncating variant had severe disease (2 DCM, 1 CPC). Whether this variant is functionally deleterious remains debated in the literature^35,95–99^, but its enrichment in severe cases in our cohort is consistent with a contributory role.

CLEC7A also harbors missense variants including p.Ile223Ser (rs16910527) which is seen in a heterozygous state in eight individuals (**Figure S11D**). The substitution replaces a hydrophobic isoleucine with a polar, uncharged serine at a residue previously shown to be required for β-glucan binding^69^. In our cohort, 6 of 8 carriers had primarily African global genetic ancestry and 87.5% (⅞) had both severe disease and African local genetic ancestry for the affected allele. The variant was significantly enriched in DCM patients (p=0.011, **Figure S11E**) and in those with African global genetic ancestry (p=0.0011, **Figure S11F**), identifying African ancestry-associated alleles to dissemination risk.

### Rare variants in *NLRX1* are enriched in individuals with DCM

The gene ontology (GO) term ‘Immune response’ was enriched in annotations for the differentially expressed genes in our RNAseq analyses (**Figure 3**, 22 genes with the annotation, FDR=3.30x10^-4^). We looked to see if there were any rare genetic variants in a curated set of 1,823 immune response genes that could explain these differences in expression (**Figure S10C**). After quality control and stringent filtering (Methods), we identified 1,327 stop-gained, frameshift, missense, and previously reported pathogenic variants across 663 immune response genes in our cohort (**Table S6**). A majority of patients (622/738) carried at least one filtered variant in an immune response gene (**Figure S10D**).

We then tested whether any immune response genes showed enrichment for rare damaging variants in DCM versus UVF using SKAT-O, a combined burden and SNP-set kernel test^100^. Rare variant burden testing identified NLRX1(NM_001282144.2) as the top gene associated with disseminated coccidioidomycosis (SKAT-O *p*=5.85×10⁻⁴, FWER threshold *p*<6.18x10^-4^ unadjusted, **Figure 4A**). Following inclusion of global ancestry PCs as covariates, the *NLRX1* association was attenuated and did not survive FWER correction (PC-adjusted *p*=0.0104 ancestry-adjusted, **Figure S12A**) consistent with the known power reduction of global ancestry adjustment at ancestry-specific loci. To determine whether the *NLRX1* signal reflects true genetic association or population stratification, we examined the local ancestry of *NLRX1* variant carriers. *NLRX1* contained two rare missense variants: p.Arg252Trp (rs145644388), a heterozygous variant found in five DCM patients, and p.Pro790Leu (rs569295450), in one patient with CPC (**Figure 4B**, **Table 2**).

**Figure 4:**
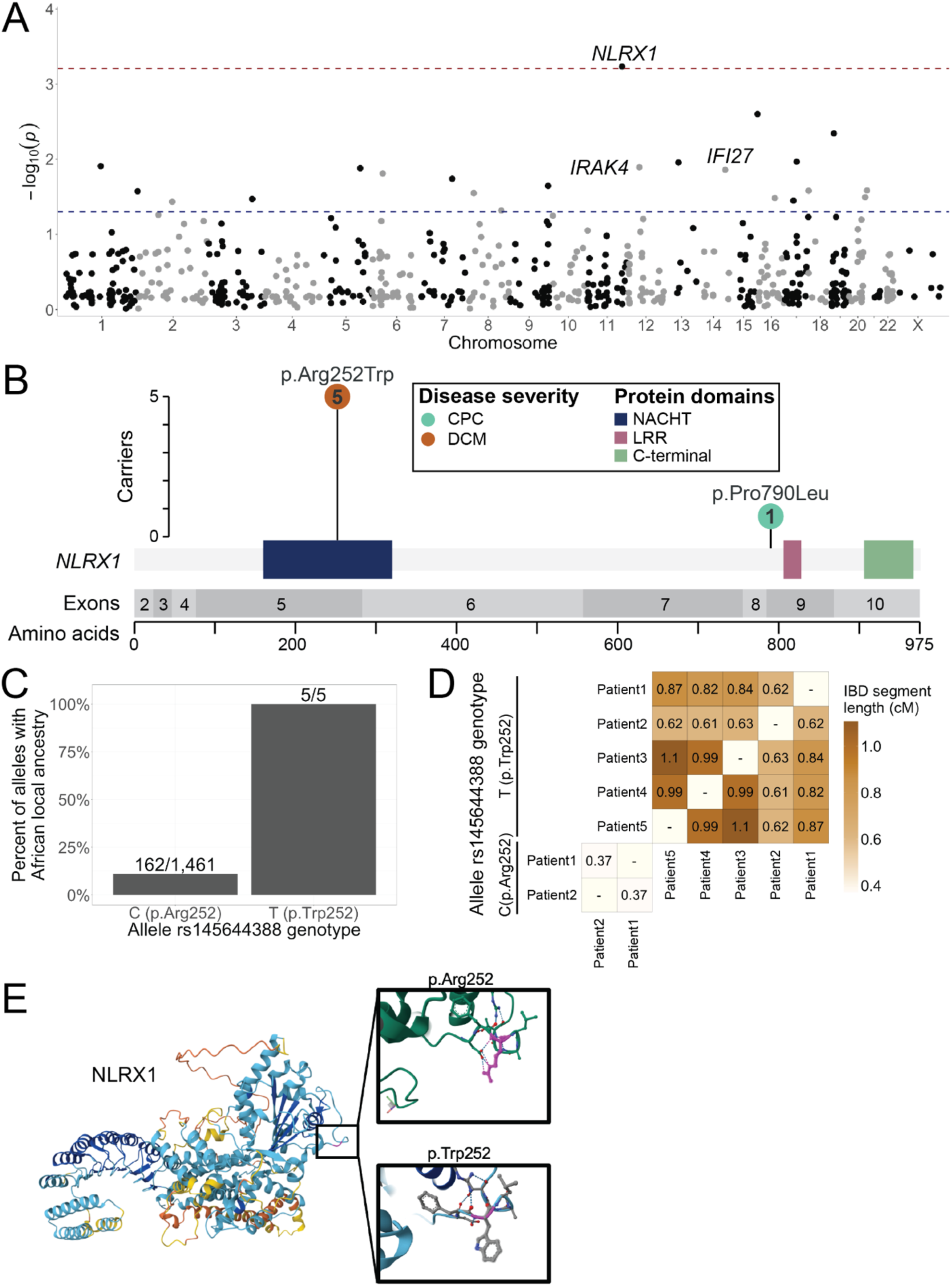
Gene-burden testing identifies *NLRX1* as a novel gene predisposing to disseminated coccidioidomycosis. (A) SKAT-O gene-based rare variant burden test results for genes in the immune response gene set. *NLRX1* was the only gene significantly enriched for rare damaging variants in DCM patients after family-wise error rate (FWER) correction with bootstrapping (*p*=6.18×10⁻⁴, red line). Nominally significant genes (*p*<0.05, blue line) with homozygous carriers (*IRAK4* and *IFI27*) are labeled. (B) Schematic of the NLRX1 protein with the locations of missense variants identified exclusively in patients with severe coccidioidomycosis (DCM or CPC). Protein domains are annotated based on UniProt. Neither variant was observed in UVF patients. (C) All alleles with the missense variant p.Arg252Trp (rs145644388, nucleotide T) share African local ancestry. Fewer than 10% of alleles without the rs145644388 variant (nucleotide C) have African local ancestry. (D) Pair-wise comparison of carrier haplotype by identity-by-descent analysis shows that carriers share between 0.6 to 1.1 centimorgans around rs145644388. The non-carrier haplotype from the same individuals (wildtype nucleotide C) at the same locus do not share this haplotype. This rare variant arose as a founder event in an African ancestry. (E) Structural modeling of p.Arg252Trp disrupts the NACHT nucleotide-binding domain in the N-terminal region that is essential for LC3-associated phagocytosis (LAP) induction. AlphaFold structural prediction indicates this variant alters local secondary structure, potentially impairing LAP-mediated fungal killing in macrophages.

**Table 2:** Rare variants potentially associated with risk of dissemination. One gene, *NLXR1*, was identified using a SKAT-O model controlling for age and sex. Seven additional rare variants that met our stringent filters (CADD Phred > 20, ESM1b or AlphaMissense predicted pathogenic) and had at least one homozygous carrier are listed. Of these, two (*IFI27* and *IRAK4*) were also nominally significant by SKAT-O testing. Using published thresholds, ESM1b scores < -7.5 and AlphaMissense scores > 0.565 were considered predicted pathogenic. Allele frequencies are from gnomAD version 4.1.1 exomes. European allele frequencies are from the non-Finish European populations. Listed are the disease severity and primary global genetic ancestry as determined by PCA for each homozygous and heterozygous carrier. ADM: Admixed American genetic ancestry, AFR: African genetic ancestry, EAS: East Asian genetic ancestry, EUR: European genetic ancestry, SAS: South Asian genetic ancestry. * “Pacific islander or Filipino” SIRE.

| Variant ID | Amino Acid Change | Severity Scores |  |  | Carriers |  | gnomAD 4.1.1 Allele Frequency |  |  | Gene SKAT-O *p*-value |
| --- | --- | --- | --- | --- | --- | --- | --- | --- | --- | --- |
|  |  | CADD Phred | ESM1b | Alpha Missense | Homozygous | Heterozygous | Overall (n=730,487) | European (n=589,658) | African (n=37,186) |  |
| rs145644388 | *NLRX1*,<br>*p.Arg252Trp* | 25.5 | -9.48 | 0.20 | 0 | 2 DCM AFR<br>2 DCM ADM<br>1 DCM SAS | 1.99x10<sup>-4</sup> | 2.25x10<sup>-5</sup> | 0.00615 | 5.85x10<sup>-4</sup> |
| rs569295450 | *NLRX1*,<br>*p.Pro790Leu* | 24.5 | -8.44 | 0.19 | 0 | 1 CPC EUR | 1.05x10<sup>-4</sup> | 1.31x10<sup>-4</sup> | 2.99x10<sup>-5</sup> | 5.85x10<sup>-4</sup> |
| rs143275071 | *IFI27*,<br>*p.Ser63Leu* | 21.8 | -7.52 | 0.29 | 1 DCM AFR | 3 DCM AFR<br>1 DCM ADM<br>1 CPC AFR | 5.12x10<sup>-4</sup> | 2.41x10<sup>-4</sup> | 0.0106 | 0.0138 |
| rs114820168 | *IRAK4*,<br>*p.Arg391Cys* | 33 | -10.77 | 0.35 | 1 DCM EAS | 2 DCM EAS*<br>1 DCM SAS*<br>1 CPC EAS<br>1 UVF EAS<br>1 UVF ADM | 1.51x10<sup>-4</sup> | 4.43x10<sup>-5</sup> | 3.00x10<sup>-5</sup> | 0.0128 |
| rs781255614 | *CR2*,<br>*p.Gly432Glu* | 23.6 | -8.11 | 0.25 | 1 DCM SAS | 1 UVF ADM | 3.17x10<sup>-4</sup> | 0 | 0 | 0.943 |
| rs201480125 | *CFHR4*,<br>*p.Trp68Ter* | 34 | -11.66 |  | 1 DCM AFR | 0 | 7.32x10<sup>-5</sup> | 5.938x10<sup>-5</sup> | 0.00105 | 0.167 |
| rs199793364 | *CCL15*,<br>*p.Thr73Met* | 24.1 | -8.79 | 0.41 | 1 CPC ADM | 1 DCM ADM<br>2 UVF ADM | 1.03x10<sup>-4</sup> | 4.05x10<sup>-5</sup> | 2.99x10<sup>-5</sup> | 0.621 |
| rs141653612 | *SEMA7A*,<br>*p.Arg109Cys* | 22.8 | -9.51 | 0.16 | 1 UVF ADM | 2 CPC ADM<br>1 CPC EUR<br>2 UVF ADM | 0.00161 | 0.00190 | 1.79x10<sup>-4</sup> | 0.108 |
| rs750740148 | *EP300*,<br>*p.Thr594Met* | 26.6 | -12.75 | 0.97 | 1 UVF EAS | 0 | 2.26x10<sup>-5</sup> | 1.71x10<sup>-5</sup> | 0 | 0.572 |

Despite heterogeneous global genetic ancestry among the five carriers of p.Arg252Trp (two dmixed American, two African, and one South Asian genetic ancestry), *NLRX1* rare variant carriers shared a common local African ancestral haplotype at the NLRX1 locus (**Figure 4C**). We next asked whether the NLRX1 p.Arg252Trp arose on an ancient founder haplotype or whether it arose independently in each distinct population. We performed pairwise identity-by-descent analysis and revealed that these carriers share between 0.6 to 1.1 centimorgans (cM) of genomic sequence around the rare variant (**Figure 4D**), consistent with a founder event approximately 45–83 generations ago (∼1,100–2,500 years)^101^. In gnomAD v4.1.1, p.Arg252Trp has a markedly higher allele frequency in African populations (AF=0.0061) compared to European populations (AF=0.00013), further supporting ancestry-specific enrichment of this variant (**Table 2**). This pattern — divergent global ancestry but convergent local ancestry — is the expected signature of a true association at an ancestry-specific locus, and is inconsistent with a spurious signal driven by global ancestry stratification *NLRX1* modulates NF-κB and type I interferon signaling, mitochondrial reactive oxygen species (ROS) production in response to bacterial and viral infection, and antifungal immune responses in macrophages^102–105^. In *Aspergillus fumigatus* infection models, NLRX1 loss increases fungal burden and pulmonary inflammation, with elevated monocyte and neutrophil numbers^106^. Both induction of LC3-associated phagocytosis (LAP) through interaction with the mitochondrial protein TUFM and changes to metabolic processing are antifungal mechanisms of *NLRX1* in macrophages^107,108^.

AlphaFold modeling predicts that the p.Arg252Trp variant disrupts the NACHT nucleotide-binding domain in the N-terminal region where TUFM interaction occurs (**Figure 4E**), potentially impairing LAP-mediated fungal containment in macrophages and reducing production of TNF, IL-6, and IL-1β^109^. While one heterozygous carrier had matched transcriptomic data and showed an increased monocyte proportion and elevated CXCL1 expression consistent with *NLRX1* loss-of-function predictions^106,109^, the limited sample size (n=1) precludes statistical testing of the significance of these associations (**Figure S13**).

### IFI27 p.Ser63Leu is an ancestry and sex-specific risk variant for severe coccidioidomycosis

Among genes nominally significant in the SKAT-O analysis, our cohort had both heterozygous and homozygous carriers of a variant in *IFI27* (**Table 2**, **Figure 4A**). *IFI27* p.Ser63Leu (rs143275071) was identified exclusively in patients with severe coccidioidomycosis — five carriers with DCM and one with CPC — suggesting a specific association with dissemination risk. *IFI27* encodes the interferon-stimulated protein ISG12, a mitochondrial protein involved in apoptotic signaling pathways critical for clearance of intracellular pathogens, including parasites like *Cryptosporidium parvum*^110,111^. Increased *IFI27* expression is associated with disease severity in several infections, including COVID-19, RSV, and leishmaniasis^82–85,112^.

All six *IFI27* p.Ser63Leu carriers are male, indicating a possible sex-specific association with this variant. In our transcriptomic analysis, *IFI27* was one of the genes significantly upregulated in male DCM patients compared to UVF (**Figure S4B**), consistent with the observation that variant carriers show dysregulation of this pathway. Five of the six *IFI27* p.Ser63Leu carriers have primarily African global genetic ancestry and six out of seven affected alleles have African local genetic ancestry. This ancestry enrichment points to a potential ancestry-driven mechanism underlying the association between genetic ancestry and DCM risk and directly links this rare variant to the 13-fold increased odds of DCM observed in individuals with African ancestry (**Figure 2**).

One *IFI27* p.Ser63Leu heterozygous carrier had matched transcriptomic data. In this patient, we observed higher *IFI27* expression (**Figure S14A**) and elevated estimated mast cell proportions (**Figures S14B–C**), consistent with previous observations that CXCL1-mediated immune responses correlate with mast cell abundance^84^. Despite these cell type and expression differences, the limited sample size (n=1 heterozygous carrier with RNA-seq) precludes statistical testing of whether these findings are significant.

## DISCUSSION

This study presents the largest comprehensively characterized cohort with laboratory confirmed coccidioidomycosis, enabling a rigorous examination of host genetic factors that modulate disease severity. Our findings corroborate and substantially extend prior epidemiological observations by demonstrating that African genetic ancestry — quantified continuously from genome sequencing rather than inferred from SIRE — is a strong risk factor for disseminated coccidioidomycosis. We further show that SIRE is an imperfect surrogate for genetic ancestry, and that associations previously attributed to SIRE categories are better explained by underlying genomic variation. Collectively, our data implicate disruption of innate immune sensing and interferon-mediated signaling as central mechanisms in the pathogenesis of severe disease and identify *NLRX1* and *IFI27* as novel candidate susceptibility genes for severe coccidioidomycosis. Notably, the NLRX1 p.Arg252Trp risk allele is found across 5 individuals spanning three global ancestry groups (AFR, ADM and SAS), but the haplotype traces back to a single ancestral founder haplotype of African origin — unifying rare variant signal under a common genetic explanation and offering a basis for elevated dissemination risk in individuals of African genetic ancestry.

### African genetic ancestry as a risk factor for dissemination

The enrichment of African genetic ancestry among DCM patients — 13-fold increased odds relative to non-African ancestry groups — is the most robust finding of this study. This association persists when ancestry is quantified continuously using ADMIXTURE rather than discrete SIRE categories and is not explained by HLA type after ancestry adjustment. These results suggest that the epidemiological associations between African American SIRE and severe coccidioidomycosis, consistently observed across over seven decades of clinical literature, at least partially reflect a biological contribution from genetic variation that differs in frequency between ancestry groups, in addition to social determinants of health.

We note that structural inequities — including differential access to health care, occupational exposure, and housing — are real and consequential drivers of health disparities and cannot be fully excluded here. However, the magnitude and specificity of the genetic ancestry association, the lack of an equivalent signal in admixed American (predominantly Latino) individuals who also face documented health inequities, and the identification of specific immune loci with higher allele frequencies in African genetic ancestry collectively argue for a biological component beyond social factors. This interpretation is consistent with prior work documenting ancestry-associated differences in type I interferon pathway activity and dendritic cell pathogen-sensing responses. Future work should integrate genomic data with socioeconomic variables to formally partition genetic and environmental contributions to dissemination risk.

### Interferon-inducible gene expression is linked to genetic ancestry and disease severity

Transcriptomic analysis identified upregulation of *IFI44*, *IFI44L*, *IFI27*, *IFIT1*, and *RSAD2* in DCM patients, genes whose elevated expression has previously been associated with African SIRE as part of a constitutively heightened interferon-stimulated gene (ISG) response^113^. Differences in expression of these genes may reflect cis-regulatory variants that are more common in individuals with African genetic ancestry, thereby linking genetic ancestry to immune gene dysregulation and, ultimately, to dissemination risk. Type I interferon signaling appears to be a central axis of dysregulation in severe coccidioidomycosis across multiple analyses: the GWAS nominally significant variant near *DPPA3* disrupts an IRF9 binding motif; *IFI44*, *IFI44L*, and *RSAD2* are canonical ISGs; and male-specific upregulation of *IFI27* and *IFIT1* further implicates this pathway. Whether interferon dysregulation is a cause or consequence of dissemination—or both—remains to be determined, but it represents a tractable target for future mechanistic and therapeutic investigation.

### NLRX1 and LC3-associated phagocytosis as a novel mechanism of dissemination risk

The identification of *NLRX1* as the only gene to reach significance in our rare-variant gene-burden test is a compelling finding. *NLRX1* is a mitochondrial NOD-like receptor with complex and context-dependent immune functions: it modulates NF-κB and type I IFN signaling, regulates mitochondrial ROS production, and — most relevant to fungal infection — facilitates LC3-associated phagocytosis (LAP) in macrophages through interaction with the mitochondrial protein TUFM. LAP is a non-canonical autophagy pathway that promotes acidification and killing of phagocytosed microorganisms, including fungi. In mouse models of invasive aspergillosis, *NLRX1* loss is associated with increased fungal burden and exaggerated pulmonary inflammation; in *Histoplasma capsulatum* infection, *NLRX1* in macrophages facilitates LAP-dependent cytokine production.

The p.Arg252Trp variant interrupts the NACHT domain in the N-terminal region of NLRX1 where TUFM binding — and thus LAP induction — takes place. Structural modeling suggests this variant disrupts local secondary structure, potentially abrogating TUFM interaction and impairing LAP, allowing *Coccidioides* to survive within macrophages and disseminate via the lymphohematogenous route. This mechanism is biologically plausible given that *Coccidioides* is known to parasitize macrophages as a vehicle for extrapulmonary spread^114,115^. Functional validation of p.Arg252Trp in primary human macrophages — including LAP assays, TUFM co-immunoprecipitation, and fungal killing assays — will be required to establish causality. The p.Pro790Leu variant in the LRR domain, carried by one CPC patient, may independently impair ROS production or oligomerization, but further work is needed.

Taken together, the *NLRX1* findings suggest that impaired LAP is one mechanism by which rare genetic variants increase dissemination risk and add coccidioidomycosis to the growing list of infections — including histoplasmosis and aspergillosis — in which *NLRX1*-mediated antifungal immunity is relevant. Pharmacological modulation of LAP or NLRX1 activity may represent a therapeutic avenue worth exploring in severe coccidioidomycosis.

### Sex differences in immune response and dissemination risk

Males comprised 64% of our cohort and were overrepresented among those with DCM, consistent with prior epidemiological data. Sex-specific transcriptomic analyses revealed non-overlapping differentially expressed gene sets in males and females, with type I and II interferon signaling genes predominantly dysregulated in males and ribosomal protein genes unexpectedly prominent among upregulated genes in females. *TLR8*’s escape from X-chromosome inactivation in T cells and monocytes may contribute to sex differences in innate immune sensing. The basis for the female-specific ribosomal protein upregulation is unclear but could reflect differences in translational regulation of immune responses or in the cell-type composition of blood at the time of sampling. These sex-specific patterns should be interpreted cautiously given the relatively small female-only sample sizes and warrant replication.

### Limitations

While our study is the largest sequenced coccidioidomycosis cohort to date, several limitations merit consideration. Because coccidioidomycosis is a rare disease, we were limited in our sample size for common variant and eQTL analyses. We estimate that as few as 25 more DCM patients would give us 80% power for our GWAS analysis (**Fig. S4C**), so continued recruitment will help further explore these factors. Because of our limited sample size, we grouped all anatomical sites of dissemination together. Bone, skin, and CNS dissemination may have distinct genetic determinants. Additionally, the blood samples used for RNA extraction were collected at the time of enrollment, meaning that patients with active disease may have different transcriptomic profiles than those who are clinically stable. Because patients with CNS DCM are more likely to have active disease than those with other forms of coccidioidomycosis, differentially expressed genes and estimated cell type proportions may reflect a heightened immune response rather than pathways unique to DCM. Our cohort lacks uninfected endemic controls, which prevented us from identifying loci associated with susceptibility to infection itself rather than severity.

While *NLRX1* p.Arg252Trp shows genetic ancestry stratified allele frequencies, several observations support a true biological association rather than confounding. First, all five carriers across three major genetic ancestry groups (African, admixed American, South Asian) presented with severe DCM, showing uniform phenotypic severity regardless of global genetic ancestry background. If the association was confounded by an association with genetic ancestry, we would expect variation in disease severity to be tied to genetic ancestry (e.g., more severe disease in carriers with African global genetic ancestry due to elevated background DCM risk in that population). Instead, the absence of ancestry-stratified phenotypic heterogeneity is consistent with a causal genetic effect. Second, the founder haplotype structure (0.6–1.1 cM IBD) indicates *NLRX1* p.Arg252Trp is a genuine segregating variant that spread through populations via migration history rather than a marker of ancestry composition. Finally, the mechanistic plausibility of *NLRX1* disruption in antifungal immunity through LC3-associated phagocytosis provides biological support for the association. However, these observations are consistent with but do not definitively prove causality. The most rigorous test — stratified burden analysis demonstrating *NLRX1* enrichment in cases vs. controls within each ancestry group — was limited by sample size (1–2 carriers per ancestry) and would require larger ancestry-stratified cohorts to fully resolve.

Finally, some of the observed genetic ancestry associations may reflect structural health inequities captured by ancestral genetic variation alone. We did not quantify health care access or socioeconomic factors, and we cannot exclude their contribution. However, the magnitude of the African genetic ancestry association and the identification of rare founder variants among 5 carriers of different global ancestries but within a shared IBD locus of local African genetic ancestry suggests that this variant arose in a single founder rather than multiple times independently in different ancestry groups.

### Conclusions

This study establishes African genetic ancestry as the strongest known risk factor for disseminated coccidioidomycosis and identifies interferon dysregulation and *NLRX1*-mediated LC3-associated phagocytosis as mechanistic contributors to severe disease. These findings move the field beyond SIRE-based associations toward a genetically grounded understanding of dissemination risk and identify specific biological pathways for therapeutic targeting. As coccidioidomycosis continues to expand in range and incidence, genomically informed risk stratification and new therapeutic strategies will be increasingly important for patients with this neglected fungal infection.

## METHODS

### Meta-analysis of coccidioidomycosis studies

Published data from seven epidemiological studies were harmonized and combined to calculate odds of dissemination in each self-identified race or ethnicity (SIRE) group compared to all other participants^21,41–43,45,116^. The labels used to describe SIRE groups and disease severity differ between publications, so classification definitions and sample counts can be found in **Table S8**. Fisher’s exact tests were used to determine the odds ratio of DCM in each SIRE group compared to all other groups using R version 4.4.1.

### Patient recruitment and classification

A dataset of 468 patients with whole exome sequencing (WES) were recruited and consented through the UC Davis IRB, as previously described in Hsu et al. 2022^35^. 299 patients were diagnosed with primary pulmonary infections, had no evidence of recurrence 2 years after discontinuation of antifungal medication, and were labeled as having uncomplicated Valley Fever (UVF). We categorized those labeled as “other” in the dataset (N=60), including those with chronic pulmonary infections that remained on an antifungal for more than a year, as having complicated pulmonary coccidioidomycosis (CPC). The remaining 109 patients with extra-pulmonary manifestations were classified as disseminated coccidioidomycosis (DCM).

An additional 329 patients were recruited from the Valley Fever Institute in Kern County, California and consented through UCLA IRB #19-001109. Using the disease severity metric described by Krogstad et al. 2023^7^, clinicians assigned each patient a severity score (**Table S9**). Those with scores of 1 or 2 were categorized as having UVF (n=115). Those with a score of 3 were categorized as CPC (n=61). Those with a score of 4, 5, or 6 were categorized as DCM (n=150). Coccidioidomycosis incidence at recruitment locations was visualized using 2021 data from the California Department of Public Health^117^.

### Patient demographic characterization

Associations between demographic variables and disease severity were evaluated using standard statistical tests. Categorical variables, including sex and sequencing type, were assessed using Pearson’s Chi-squared test. Continuous variables, such as age and proportion of African genetic ancestry, were analyzed using the Kruskal-Wallis rank sum test. Fisher’s exact test was used for categorical count data, including self-identified race/ethnicity (SIRE) and major genetic ancestry. To contextualize our cohort’s diversity, SIRE and genetic ancestry distributions were compared to population estimates from the 2020 California Census^46^.

### DNA sequencing and processing

Whole exome sequencing (WES) of the UC Davis participants was conducted as described in Hsu et al. 2023^35^. Whole genome sequencing (WGS) of the other patients was performed using a NovaSeq6000 to 27-30X coverage and processed using Illumina DRAGEN pipelines.

### Exome LiftOver and genotype imputation

Since the UC Davis data was originally aligned to the hg19 human reference and the raw files were not available to us, we used UCSC LiftOver to convert the variant positions to hg38. We converted the variant call format (VCF) files to PLINK 1.9 format and extracted the chromosome and position information for each variant to feed into the web version of the software^118,119^. Using a custom R script, we created lists of the variants that were successfully lifted over and those that could not be lifted over. With PLINK we removed the variants with no equivalent position in hg38 and updated the positions of all other variants. We also removed variants from non-standard chromosomes.

We then prepared the data for imputation with the TOPMed imputation server^120,121^. We first used PLINK to calculate the variant allele frequencies. Then, a reference panel for preprocessing was constructed from the full list of TOPMed variants that have been submitted to dbSNP as described by Dr. William Rayner^122,123^. Because few of our rare variants were found in this reference panel, variants with MAF < 0.01 were filtered. We used the resulting dataset and reference panel to filter and reformat our PLINK files into per chromosome VCFs using Dr. Rayner’s preprocessing script^123^. Chromosome names were edited to include the “chr” prefix. The VCFs were then zipped and indexed with bcftools. The resulting per chromosome VCFs were uploaded to the TOPMed imputation server, which phased and imputed the data^124,125^. Resulting imputed VCFs were downloaded, unzipped, and filtered to only high-quality variants (imputation R^2^ > 0.3). All variants present in the original dataset that were not included in the final imputed VCFs were identified and added to the VCFs with BCFtools *isec* and *concat* commands^126^. Since PLINK 1.9 changes the allele orders, we used the BCFtools *norm* command to match the reference and alternate alleles to the hg38 reference sequence.

### Data merging

In order to include all possible variants in our final dataset, all WGS data was combined as genomic VCFs (gVCFs). Using the BCFtools *annotate* command, we removed the allele frequency (AF) tag from the first WGS batch to enable merging with later batches. We then merged the batch gVCF files by chromosome and indexed the resulting gVCFs using the BCFtools *merge* and *index* commands. Although gVCFs were not available for the WES data, the large sample size (n=468) meant that there were still many rare variants included in the VCFs. The WES per-chromosome VCFs were combined with the merged WGS per-chromosome gVCFs and indexed using BCFtools *merge* and *index* commands. The resulting WES and WGS gVCFs had the variant INFO and FORMAT fields recalculated using the BCFtools *+fill-tags* add-on and were converted to VCFs using BCFtools *view* options. All per-chromosome VCFs were then combined into one VCF file using BCFtools *concat*.

Using the BCFtools *view* and *filter* commands, we kept only variants with passing FILTER fields, read depths (DP) more than 10, and genotype qualities (GQ) more than 20. We removed all duplicated variants and standardized allele order with BCFtools *norm --rm-dup ‘all’*. We calculated the missingness, singletons, and depth for each patient using vcftools. We then carried out standard PLINK quality control steps with PLINK 1.9 and PLINK 2.0^127^. After converting the VCF files to PLINK format, we added the sample ID, phenotype, and sex information and standardized variant IDs. This dataset included 35,984,100 variants from 760 samples.

### Common variant quality control

For genetic ancestry and common variant analyses, we removed variants that were not in Hardy-Weinberg equilibrium (HWE, *p*<0.0001), variants with more than 10% of patients with missing genotypes (GENO), and patients with more than 10% of genotypes missing across all variants (MIND, n=18). We removed one of each pair of first-degree relatives (kinship threshold > 0.177, n=9) using the PLINK implementation of Manichaikul et al.’s KING robust estimator^128^. We then filtered to biallelic variants with a minor allele frequency (MAF) of at least 0.05 in our sample. Linkage disequilibrium (LD) pruning was carried out for variants with a pairwise R^2^ > 0.1. This LD pruned data was used to check for mismatches in genetic sex with reported sex using the PLINK --check-sex ycount option. After removing samples with mismatched sexes (n=8) from the dataset without LD filtering, we converted the dataset back to a VCF and again used the BCFtools +fill-tags add-on to recalculate INFO and FORMAT fields removed by PLINK. The final QC-passed VCF contained 1,001,289 variants from 733 patients. We also removed samples with mismatched sexes from the PLINK LD-pruned dataset to create a final PLINK dataset for global genetic ancestry calculation. This final LD-pruned PLINK dataset contained only 419,990 variants.

### Global genetic ancestry calculation with ADMIXTURE

We used the phased, harmonized data from the Simons Genome Diversity Project (SGDP), the Human Genome Diversity Project (HGDP), and the 1000 Genomes Project (1000G) as a reference for genetic ancestry calculation and phasing^129–132^. After merging these 3,356 reference samples with the filtered LD-pruned PLINK dataset, we ran an unsupervised model with k=5 using ADMIXTURE 1.3.0 to estimate global genetic ancestry proportions^50^. By comparing the proportions of each unsupervised group to the population labels of the reference samples, we determined that population 1 was East Asian, population 2 admixed American, population 3 European, population 4 South Asian, and population 5 African genetic ancestry. We compared the proportion of variants estimated to come from each genetic ancestry to patient coccidioidomycosis severity with a *t*-test. The total number of heterozygous variants per patient was calculated with the PLINK 2.0 --sample-counts method, which was then compared with the proportion of predicted African genetic ancestry using a linear regression.

### Principal components analysis

Before conducting principal components analysis (PCA) on the merged PLINK dataset, we removed all reference samples from diaspora populations (e.g. STU, Sri Lankan Tamil in the UK) and from the Middle East and Papua New Guinea in order to more easily separate patients into discrete groups (see **Table S10**). This left us with a reference dataset of 2,546 samples. We then updated the sex and set the phenotype to 1 for all reference and patient samples to enable running the desired model with EIGENSOFT’s SMARTPCA^133,134^. Using a custom R script, we visualized the calculated PCs and chose thresholds that separated the reference samples into five ancestral groups using the first four PCs (**Figure S1B**). Patient samples with PC1 > 0.01 were designated as having African genetic ancestry. Those with PC3 > 0 were considered to have admixed American genetic ancestry. Those with PC2 > 0.01 were considered to have East Asian genetic ancestry, and those with PC2 > -0.015 or PC4 > 0.02 were designated as having South Asian genetic ancestry. All remaining samples clustered best with European genetic ancestry reference samples.

### Common variant phasing and local ancestry calling

Using the phased reference dataset chromosome files without filtering, we used SHAPEIT 5.1.1 to phase the common variant QC-passed VCF files^135^. Then, we used the filtered reference dataset chromosome BCFs and the assigned population labels as the reference and sample map to call local ancestry for the phased patient BCFs using RFMix v2.03^136^.

### Genome wide association tests

Using the QC-passed common variant PLINK files, we ran a PLINK2 logistic regression model comparing dissemination status (UVF and CPC vs DCM) to genotype, controlling for age, sex, and the first ten PCs generated by PLINK. We used the “--covar-variance-standardize” option to transform the quantitative covariates to mean 0, variance 1. A standard GWAS threshold (α = 5x10^-8^) was used to assess statistical significance. We considered associations nominally significant if they had p-values less than 1x10^-5^. We annotated nominally significant variants using HaploReg and Ensembl’s Variant Effect Predictor (VEP)^137,138^.

We also used the python package admix-kit, as described in Hou et al. 2024, to conduct ancestry-aware genome-wide association tests controlling for global and local ancestries^47^. However, all models implemented by this package, including admixture mapping, had inflated *p*-values.

### Power calculation

We estimated our power to detect genotype associations with our current dataset with the University of Michigan Genetic Association Study (GAS) Power Calculator^139^. We set the significance level to the standard GWAS threshold (5x10^-8^) and disease prevalence to 0.01. We set the allele frequency and genotype relative risk to the allele frequency and odds ratio of the SNP with the lowest *p*-value (AF=0.2, OR=2.2).

### HLA calling and statistical analysis

Human leukocyte antigen (HLA) types were called for the genes *A*, *B*, *C*, *E*, *F*, *G*, *DQA1*, *DQB1*, *DPA1*, *DPB1*, *DRB1*, *DRB3*, and *DRB4* with the HLA*LA software^140^ for 669 samples with available BAM files. We removed samples with missing phenotype information (n=21), who had discordant sex or were duplicate samples (n=20), and who had a mean call quality (Q1) less than 0.8 (n=2). We also removed both alleles of a gene for a sample with at least one allele call with Q1 < 0.8 or for which the algorithm had multiple possible alleles. After quality control, there were 626 samples with QC-passed HLA calls (349 UVF, 94 CPC, and 183 DCM). For each two-field HLA allele (for example, DPA1*01:03), we calculated the allele frequency in our HLA-typed patients by disease severity and graphed the allele frequency differences. Because we were primarily interested in Type II MHC genes and HLA*LA has not been optimized for calling DRB3 or DRB4 alleles, we looked at results for DPA1, DPB1, DQA1, DQB1, and DRB1 alleles only. We used Fisher’s Exact Tests as an initial filter to test for correlations between having an allele and disease severity. We used a threshold of *p* < 0.0004 because we tested 124 alleles from the five genes of interest.

For alleles with AF >= 0.05, we then used a logistic regression model controlling for age, sex, sequencing batch, and PCA global genetic ancestry, to more rigorously test for association between disease severity and having the allele. This model was chosen because the *p*-values generated were normally distributed (**Figure S5D**). We adjusted for multiple testing across all DPA1, DPB1, DQA1, DQB1, and DRB1 alleles with the Benjamini and Hochberg false discovery rate (FDR) method^141^.

### RNA-sequencing and quality control

RNA sequencing was carried out for 276 patients across six sequencing runs using the QIAseq FastSelect -rRNA HMR Kits with NEBNext® Ultra™ II RNA Library Prep Kit for Illumina® and sequenced on a NextSeq2000 with 150 bp paired-end. We removed adapter content and trimmed base pairs with quality less than 25 at the ends of reads using BBMap^142^. Reads were aligned to GRCh38 with STAR^143^ and read counts generated with featureCounts^144^. We checked whether sample genetic sex and reported sex matched by calculating the ratio of read counts of Y-chromosome genes to those of X-chromosome genes. We removed three male patients with ratios < 0.01 and three female patients with ratios > 0.01. In addition to the six samples that failed sex check, one sample with no known exposure to Coccidioides was excluded from our analyses.

Because of strong differences in gene expression between sequencing batches, we used ComBat-seq^145^ to remove batch effects, with disease severity as the biological covariate. We then removed two samples with a library size below 1 million reads and filtered our dataset to 26,460 expressed genes using the default edgeR parameters (CPM>=0.896 in at least 32 samples with at least 15 reads across all samples)^146^. In order to include age as a parameter in our differential expression model, we scaled and centered age for all patients. After quality control, we had expression information for 26,460 genes in 267 samples: 100 UVF, 45 CPC, and 122 DCM.

### Differential expression analyses

We then performed differential expression analysis between the three disease severity levels (UVF, CPC, and DCM) with DESeq2^147^, controlling for sex and age. For each comparison - UVF vs DCM, UVF vs CPC, and CPC vs DCM - we filtered to genes with an adjusted p-value less than 0.05. We also ran DESeq2 models controlling only for age in male and female patients alone. **Table S3** is a list of all genes significantly differentially expressed in any comparison.

### Protein interaction and functional enrichment analyses

Using the STRING database, we retrieved annotations for all differentially expressed (DE) protein-coding genes from the Gene Ontology (GO), Human Phenotype Ontology, Human Disease Ontology, KEGG, Reactome, InterPro, SMART, and UniProt databases, as well as several other web-scraped resources (**Table S4**)^148–159^

We curated a set of immune function terms that included the names of immune cells (e.g. “Leukocyte”, “Neutrophil”, “Macrophage”), immune signaling pathways (“JAK-STAT”, “cytokine”, “interferon”, etc.), or antigens (“virus”, “SARS-CoV”, “fungus”, etc.) as well as more general descriptors like “immune”, “defense”, and “infection” (**Table S4**, Immune annotation). We added to this list any gene listed in one of three recent publications about variants causing inborn errors of immunity (**Table S11**, IEI)^55–57^ to create our set of “immune genes” (**Table S3**, Immune gene).

We then carried out functional enrichment analysis of the nine protein-coding genes that were DE in DCM compared to UVF in the full dataset. We graphed the proportion of DE term genes for the significant GO Process and UniProt keywords terms. We also carried out functional enrichment analysis for the 86 protein-coding genes that were DE in any comparison in the full cohort, in just males, and in just females.

Finally, we created protein-protein interaction networks with medium confidence (interaction score >= 0.4) for the nine protein-coding genes with DE in DCM vs UVF patients and 86 protein-coding genes with DE in any comparison. We exported the resulting networks to Cytoscape for visualization^160^.

### Cell type proportion estimation

We wrote out our raw RNAseq readcount data to a tab-delimited file to input into the CIBERSORTx web portal^161^. Using the “Impute Cell Fractions” module and our input file as the mixture file, we used the previously calculated leukocyte signature matrix (LM22) and source GEP to run immune cell type deconvolution with B-mode batch correction^162,163^. We used absolute mode to compare our proportions across individuals and did 1,000 permutations for significance analysis. For visualization, we converted absolute cell type estimates to proportions by dividing by the absolute score (equivalent to the sum of all cell type values). We also grouped cell types into ten categories: B-cells, dendritic cells, eosinophils, macrophages, mast cells, monocytes, neutrophils, natural killer cells, plasma cells, and T-cells. We used *t*-tests to compare the proportion of each cell type in those with DCM to UVF, CPC to UVF, and DCM to CPC. We also used *t*-tests to compare the proportion of each cell type in males to females. Based on these results, we ran linear regressions correcting for sex, age, and RNAseq batch in B-cells and T-cells.

### Previously described coccidioidomycosis risk variants

We carried out a literature review of genetic studies of human host susceptibility to coccidioidomycosis and compiled a list of 36 variants described in at least one publication as being associated with disease^2,35–39,94^. We extracted genotypes for all variants available in our DNA sequencing. After filtering for variants that had fewer than 10% missing genotypes (GENO < 0.1) in Hardy-Weinberg equilibrium (HWE *p*<1x10^-8^) and samples with concordant sex missing fewer than 10% of the remaining variants (MIND < 0.1), we had 15 risk variants with genotypes in 728 samples. We annotated the variants with VEP and calculated the allele frequencies by disease severity and PCA-based genetic ancestry in our cohort (**Table S5**). We used Protter, AlphaFold, and Mol* to visualize the variants in *CLEC7A* (Dectin-1)^164–166^. We used amino acids identified by Brown et al. 2007 and Adachi et al. 2004 to be associated with β-glucan binding to annotate our Protter diagram (**Figure S11A**)^69,167^.

### Rare variant filtering and quality control

We generated a list of 1,823 genes of interest by querying the Gene Ontology (GO) database for the term “Immune response” (GO:0006955) and determined gene position using BioMart^168^ to access Ensembl Genes 115 GRCh38 positions. We then extracted all variants in those genes from the full PLINK dataset (n=760) and exported the resulting 775,854 variants to VCF format. Using VEP, we annotated the VCF for coding consequence, NCBI ClinVar pathogenicity predictions, and gnomAD 2.1.1 population allele frequencies (AF). We focused on variants with stop-gained, stop-lost, frameshift, indel, missense, and other serious coding consequences, as well as any other variant that had a “pathogenic” annotation in ClinVar. We then filtered for variants that had AF < 0.01 in all gnomAD 2.1.1 populations to focus on very rare variants. The resulting 10,671 rare variants then underwent quality control similar to that used for common variants: we kept 5,388 variants from 738 samples with GENO < 0.1, HWE *p*<1x10^-8^, MIND < 0.1, and concordant sex.

These variants were annotated further with VEP to get CADD Phred, ESM1b, and AlphaMissense scores, as well as nonsense mediated decay (NMD) predictions. We then curated a list of variants likely to have more severe consequences, all of which had either CADD Phred score > 20 or a pathogenic annotation in ClinVar: stop-gained or frameshift variants with predicted NMD, missense variants with ESM1b LLR < -7.5 or AlphaMissense score > 0.564, and variants in ClinVar with no benign annotations. The final list of filtered variants included 1,327 variants with 622 carriers (See **Table S6** and **Figure S9**).

### Gene burden testing

We used SKAT-O, a combined burden and SNP-set kernel test, on the list of 1,327 variants to see if any of the 664 genes they came from had more variants than would be expected by chance (**Table S7**)^100^. For autosomal genes, we used the SKAT_Null_Model function with 1,000 resampling iterations to generate the null model and SKATBinary function with 2x10^-6^ resampling iterations to get the model result. For genes on the X chromosome, we used the SKAT_Null_Model_Chr_X function with 1,000 resampling interactions to generate the null model and SKAT_ChrX function to get the model result. Using the resampled *p*-values from the null models across all genes, we used the Resampling_FWER_1 function to implement the package’s family-wise error rate (FWER) method, resulting in a significant *p*-value cut-off of 6.18x10^-4^ at 5% of the bootstrapped p-values for the model correcting only for age and sex.

Since genetic ancestry is associated with DCM prevalence, we considered whether ancestry adjustment was appropriate for this rare variant burden analysis. We compared SKAT-O results without (**Figure S12B**) and with (**Figure S12C**) correction for the first ten principal components using quantile-quantile (Q-Q) plots. Correcting for genetic ancestry PCs showed deflation of all *p*-values, consistent with removal of ancestry stratification (**Table S7**).

### Rare variant phasing

We used the phased common variants VCF as a scaffold, in addition to the phased reference dataset chromosome files, to phase rare variants from immune response genes using the SHAPEIT 5.1.1 phase_common method. This is appropriate because of our small sample size (< 2,000 samples)^135,169^. We used the phased genotypes to determine the local genetic ancestry of the haplotype a rare variant of interest fell on.

### Identity by descent

Using the software package phase-ibd, we detected regions of shared identity-by-descent (IBD) that shared at least 5 SNPs in a 0.05 cM region (min-seed=0.05, min-extend=0.01, min-output=0.05, min-markers=5). This allowed us to detect regions of IBD shared between rare variant carriers. We quantified and plotted the length of the largest shared IBD segment on the variant and non-variant allele haplotypes.

### Data and Code availability

All scripts used to carry out quality control and analysis are included on the GitHub repository https://github.com/Spendlove/cocci.

## Data Availability

All data produced in the present study are available upon reasonable request to the authors. The whole genome sequencing data and the transcriptomics data will be made available via dbGAP upon publication of the manuscript.

## Acknowledgments

We thank the patients who participated in our study. We would like to thank Smriti S Nagarajan and Miguel A Moreno Lastre for processing samples. Finally, we would like to thank the members of the Arboleda Lab, Butte lab, and Pasaniuc lab for their guidance throughout the manuscript and the tireless participation of patients and families in our project that make this work possible. Some of the figures were created in https://BioRender.com

## Contributions

**[SLJ]:** Investigation, Writing-original draft, review & editing, **[SJS]:** Investigation, Writing-review & editing, **[AVS]:** Resources, Data Curation, **[ZJ]:** Investigation, Formal analysis, **[VA]:** Methodology, **[KH]:** Methodology, **[RM]:** Methodology, **[TT]:** Methodology, **[EE]:** Resources **[GRT]:** Resources, Data Curation, **[RHJ]:** Resources, Data Curation, [**AH]:** Resources, Data Curation, **[RK]:** Resources, Data Curation, **[BP]:** Conceptualization, Funding acquisition, supervision, **[HP]:** Data Curation, Review and editing **[PK]:** Data Curation, Review and editing. **[MB]:** Funding acquisition, supervision, Writing-review & editing, **[VAA]:** Funding acquisition, supervision, Writing-original draft, review & editing

