## Supplemental Files for "Genetic and transcriptomic determinants of disseminated coccidioidomycosis identify a founder variant in *NLRX1* and ancestry-specific rare variants in immune response genes"

Supplemental Figures related to Figure 1....................1
Supplemental Figures related to Figure 2....................2
Supplemental Figures related to common variants analysis.................... 4
Supplemental Figures related to Figure 3....................6
Supplemental Figures related to Figure 4....................13

### Supplemental Figures related to Figure 1

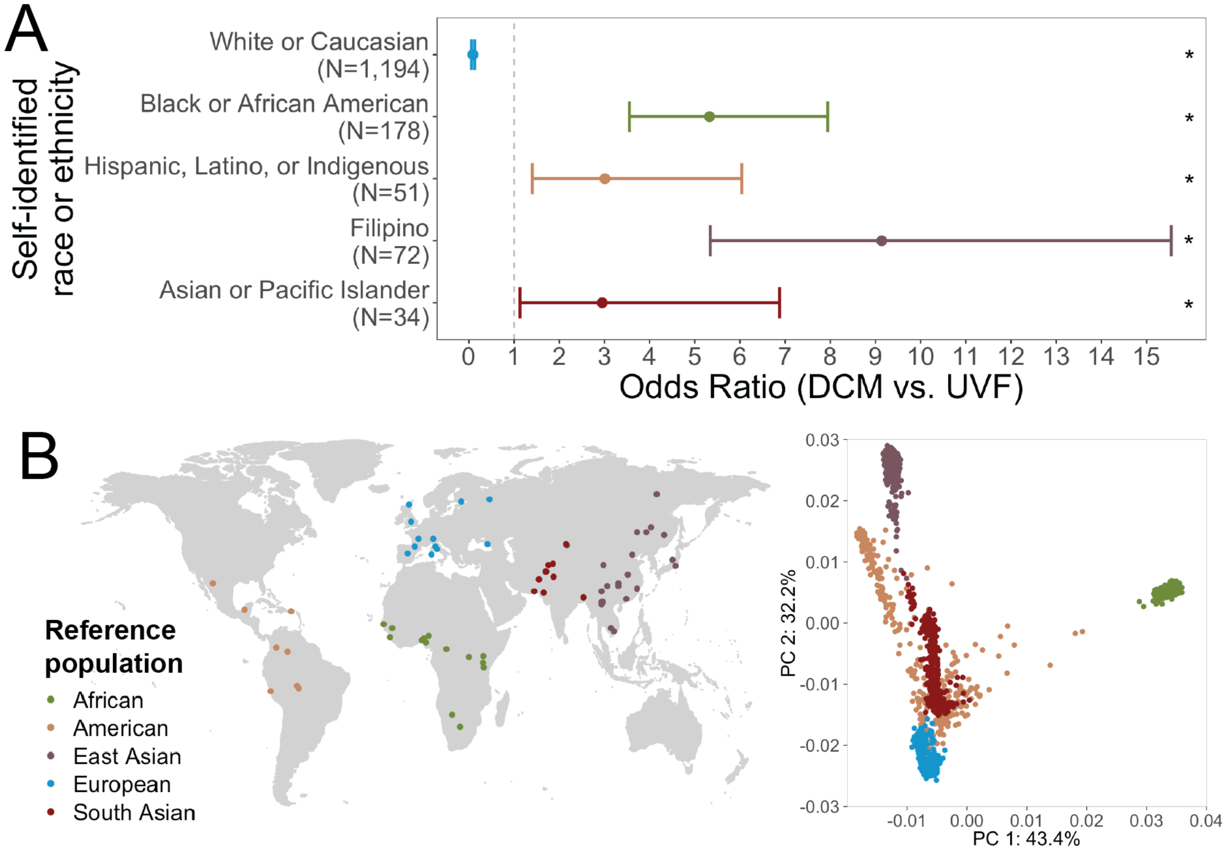


**Figure S1: Self-identified race/ethnicity (SIRE) and genetic ancestry are each associated with risk of disseminated coccidioidomycosis (DCM).**

**(A)** Meta-analysis of six epidemiological studies published between 1944 and 2008 shows that individuals who self-identified as a non-European race or ethnicity had significantly higher odds of dissemination (Black: OR = 5.33, $p = 1.26\times10^{-15}$; Latino: OR = 3.01, $p = 2.60\times10^{-3}$; Filipino: OR = 9.14, $p = 1.96\times10^{-15}$; Asian or Pacific Islander: OR = 2.95, $p = 0.014$). White subjects were significantly less likely than all other groups to have DCM (OR = 0.09, $p = 1.59\times10^{-40}$). All comparisons were calculated with Fisher's Exact Test and are significant (marked with asterisk). **(B)** To quantify genetic ancestry, we performed principal components analysis (PCA) using PLINK with reference samples (n = 3,356) from the 1000 Genomes Project (1000G), Human Genome Diversity Project (HGDP), and Simons Genome Diversity Project (SGDP). Collection sites for reference samples are shown on the world map (left), with colors indicating continental ancestry group. Clustering of reference samples along PCs 1–4 (right) was used to define ancestry boundaries for each primary population group. African genetic ancestry (green) was determined based on PC1.

#### Supplemental Figures related to Figure 2

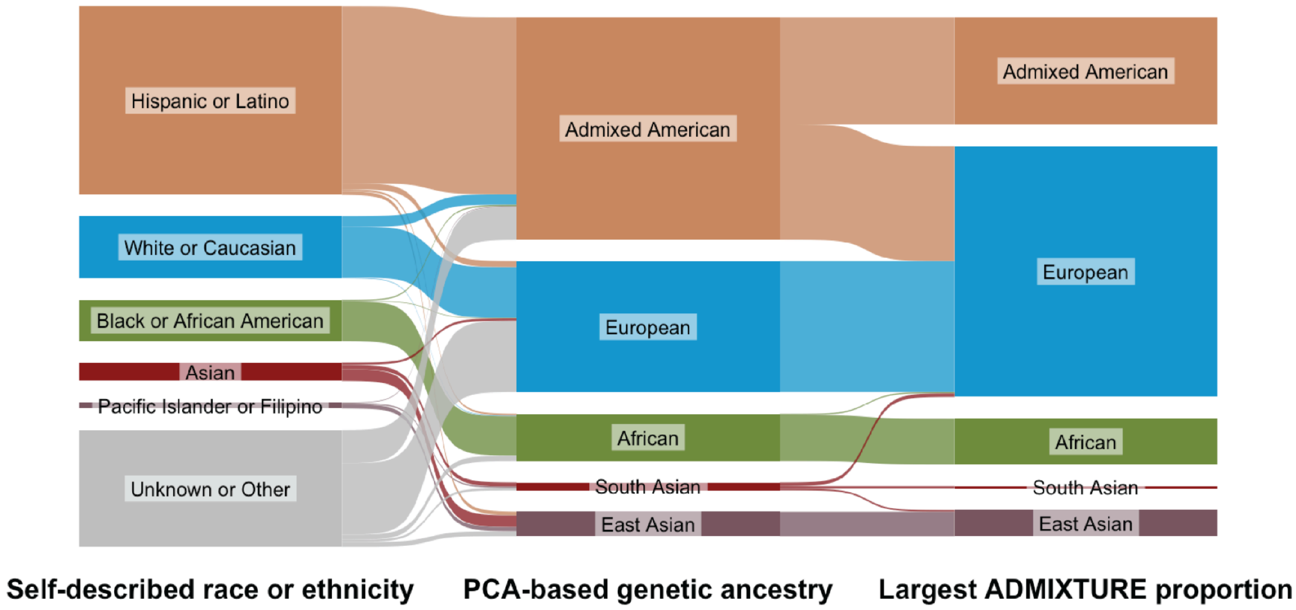


**Figure S2: SIRE and genetic ancestry classifications are not always concordant.**

Self-identified race or ethnicity (SIRE) is a social construct that is not always reflected at the genomic level. Several individuals who self-identified as non-White clustered with European reference samples in PCA, including four with "Asian" SIRE, one with "Black or African American" SIRE, and ten with "Hispanic or Latino" SIRE. Although individuals with Latino heritage often self-identify as "White or Caucasian," 18 subjects in our cohort who did so clustered with indigenous American reference populations (labeled "Admixed American") in PCA. We performed unsupervised ADMIXTURE analysis combining our data with five continental reference groups to estimate the proportion of each sample's DNA derived from each of five ancestral populations. Although the same reference populations were used for both PCA-based and ADMIXTURE-based ancestry assignments, these two methods did not always agree. For example, because many individuals with "Hispanic or Latino" SIRE carry both European and indigenous American ancestry, and because ADMIXTURE-based proportions vary continuously, approximately half of these individuals had a higher estimated proportion of Admixed American ancestry and half had a higher estimated proportion of European ancestry, even though all clustered together in the PCA.

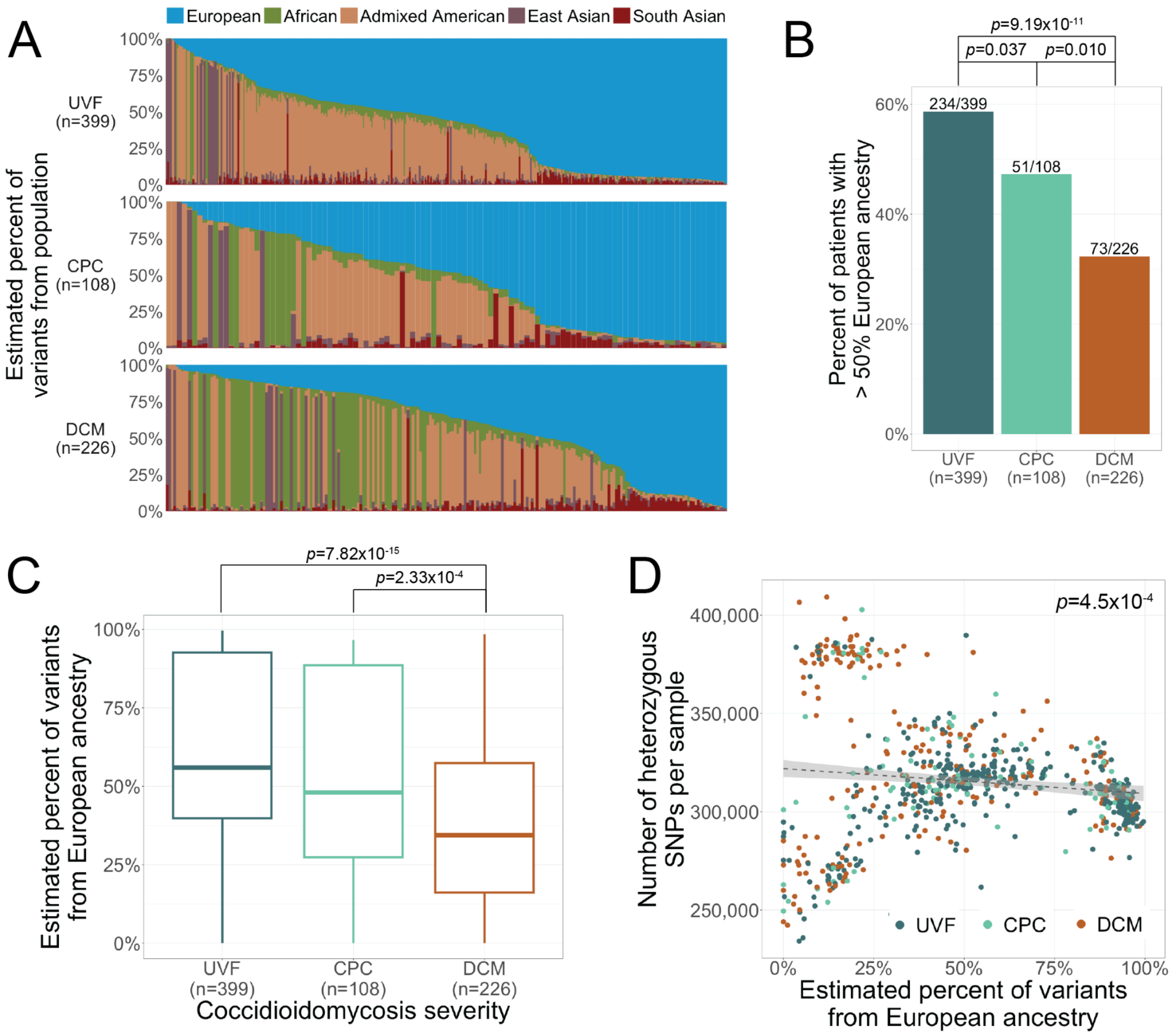


**Figure S3: European genetic ancestry is associated with significantly lower risk of dissemination.**

(A) Proportions of continental genetic ancestry estimated using unsupervised ADMIXTURE modeling (k = 5) are significantly associated with coccidioidomycosis disease severity. Individuals with high proportions of European ancestry were most likely to have uncomplicated valley fever (UVF). (B) Among UVF patients, 58.6% had more than 50% European genetic ancestry — significantly more than among DCM patients (32.3%, $p = 9.1\times10^{-11}$) or CPC patients (47.2%, $p = 0.04$) by Welch two-sample *t*-test. (C) The mean proportion of European genetic ancestry was 40.4% in DCM patients, compared with 53.8% in CPC patients ($p = 2.3\times10^{-4}$) and 59.7% in UVF patients ($p = 7.8\times10^{-15}$), as determined by Welch two-sample *t*-test. (D) Increased proportion of European ancestry was associated with decreased genetic diversity; each additional 1% of European ancestry corresponded to an estimated 127 fewer heterozygous SNPs ($p = 4.5\times10^{-4}$, linear regression).

Supplemental Figures related to common variants analysis

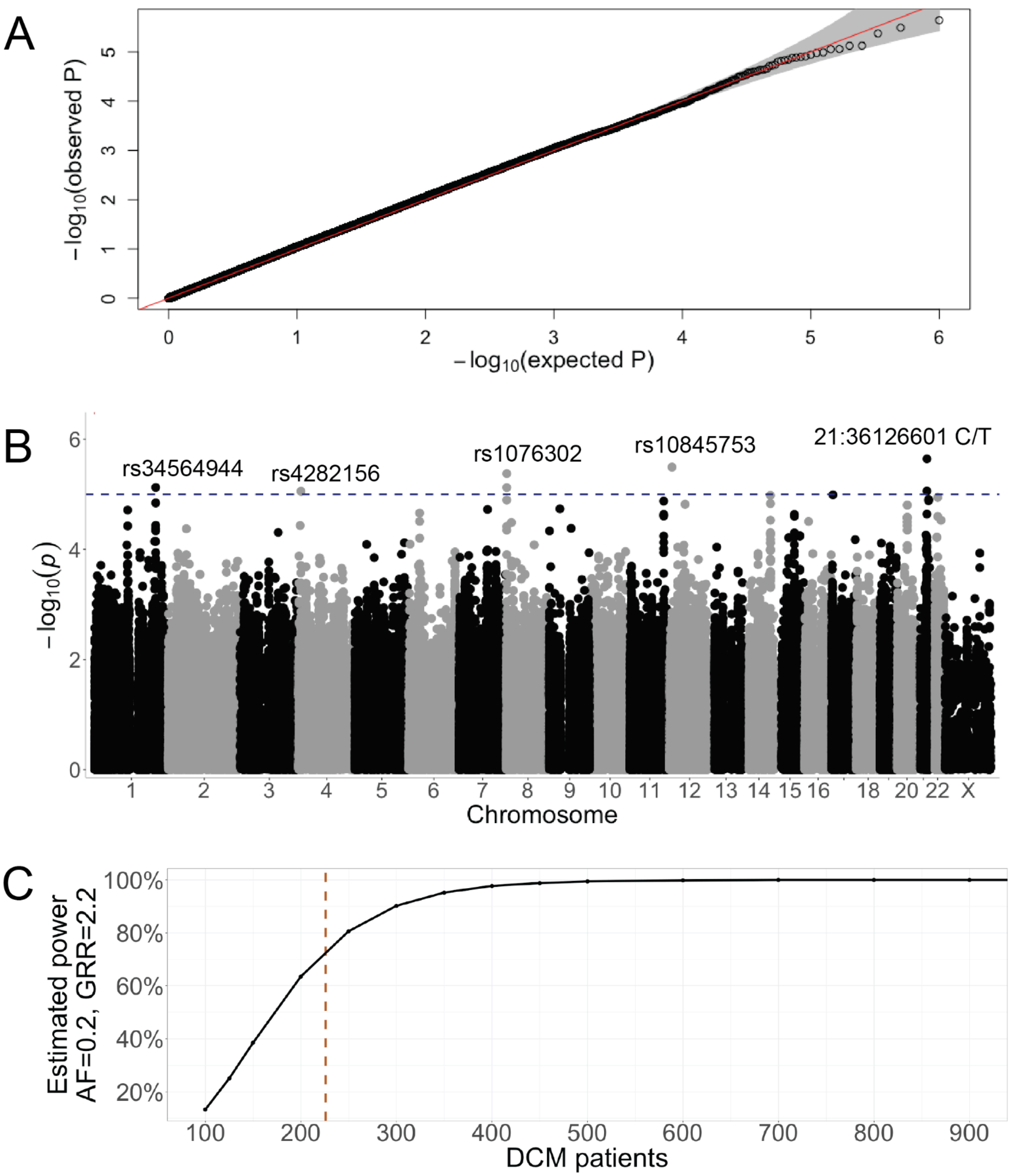


**Figure S4: Genome-wide association analysis of common variants.**

**(A)** Q-Q plot of observed versus expected *p*-values for all tested SNPs. The absence of early deviation from the null diagonal indicates that the GWAS model is well-calibrated with no evidence of genomic inflation. **(B)** Manhattan plot of GWAS results showing *p*-values for SNPs across all chromosomes. Lead SNPs for each of the five regions with nominally significant associations ($p < 1 \times 10^{-5}$, blue line) are labeled. Both nominally significant chromosome 21 variants were novel and lacked a dbSNP identifier. **(C)** Power analysis for our sample of 733 individuals with 226 DCM patients (red line) indicates 75% power to detect rare variants with a genotype relative risk (GRR) equal to the odds ratio of the SNP with the lowest *p*-value (21:36126601 C/T, OR = 2.2, $p = 2.27 \times 10^{-6}$). Given the allele frequency of this SNP (0.19), an allele frequency of 0.2 was used for power calculations. We estimate that 80% power would be achieved with an additional 24 DCM patients.

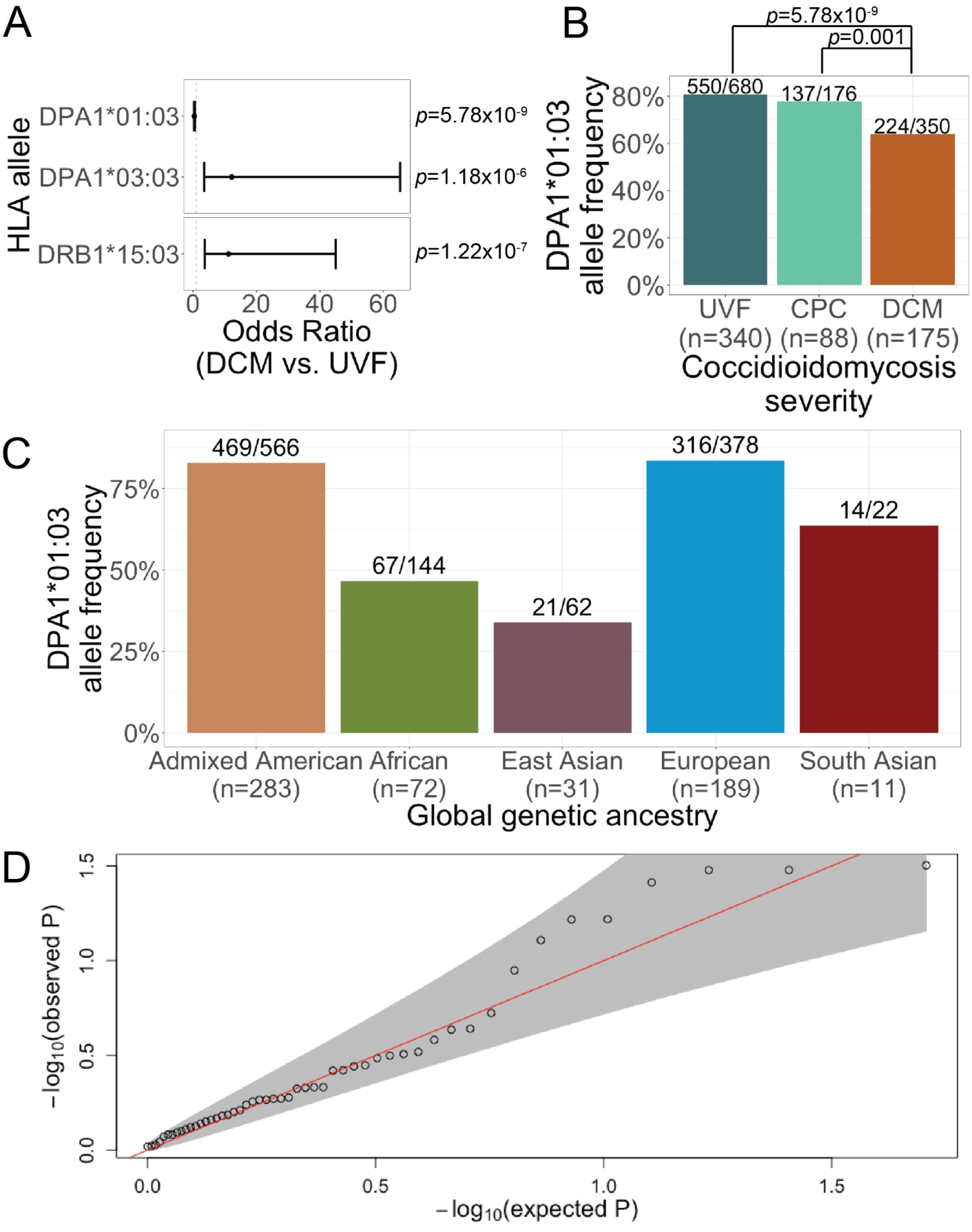


**Figure S5: HLA allele associations with coccidioidomycosis disease severity.**

(A) The *DPA1*01:03* allele was significantly less common in DCM patients than in UVF patients ($p$ = 5.78×$10^{-9}$, Fisher's Exact Test). The *DPA1*03:03* and *DRB1*15:03* alleles were each significantly more common in DCM than in UVF patients ($p$ = 1.18×$10^{-6}$ and $p$ = 1.22×$10^{-7}$, respectively). (B) The *DPA1*01:03* allele was also significantly less common in DCM patients than in CPC patients ($p$ = 0.001). The ratio above each bar represents the number of *DPA1*01:03* alleles divided by the total number of called *DPA1* alleles for each severity group. (C) *DPA1*01:03* is most prevalent in individuals with Admixed American and European genetic ancestry in our cohort. The ratio above each bar represents the number of *DPA1*01:03* alleles divided by the total number of called *DPA1* alleles for each genetic ancestry group. (D) Q-Q plot demonstrating that a logistic regression model of disease severity regressed on HLA allele presence, with covariates for age, sex, sequencing batch, and PCA-derived global genetic ancestry, does not produce inflated $p$-values.

Supplemental Figures related to Figure 3

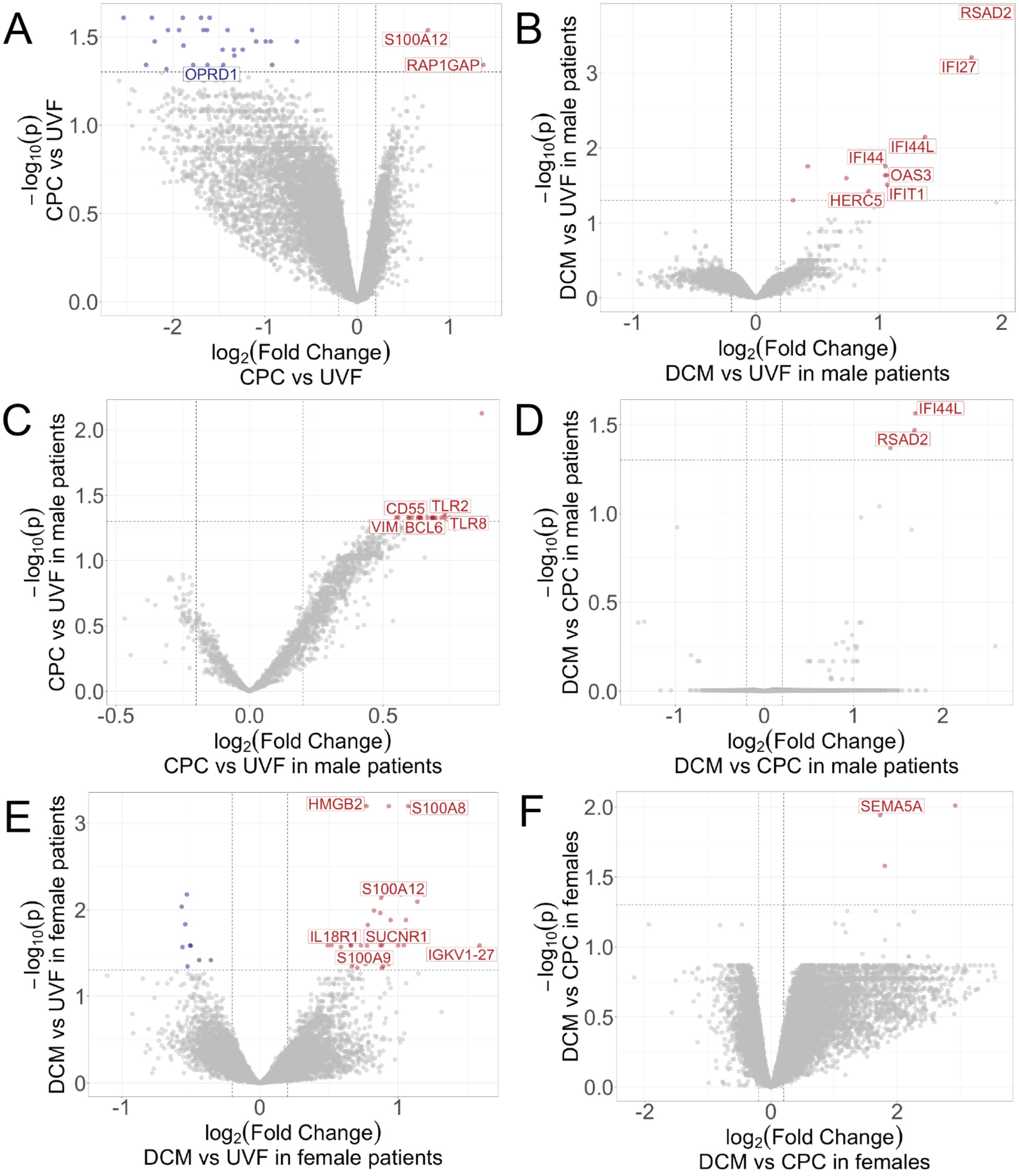


**Figure S6: Differential gene expression results across all pairwise severity comparisons.**

Volcano plots showing differential gene expression from blood RNA-seq data across 267 coccidioidomycosis patients, stratified by comparison group. Differentially expressed (DE) genes are colored by direction of effect (downregulated in blue, upregulated in red). In panels A–E, significantly DE genes annotated with the Gene Ontology (GO) Biological Process term "immune response" are labeled. **(A**) In the sex-combined cohort, 31 genes were DE between CPC patients (n = 45) and UVF patients (n = 100). All but two were downregulated in CPC relative to UVF. *S100A12* was upregulated in both CPC and DCM compared to UVF ($p = 0.03$ and $p = 0.02$, respectively). Both upregulated DE genes and 16 of 29 downregulated DE genes carried immune annotations. **(B)** In male patients, 13 genes were upregulated in DCM (n = 90) compared to UVF (n = 50), 11 of which carried immune annotations. (C) In male patients, 22 genes were DE between CPC (n = 28) and UVF (n = 50); all but 4 were immune-related. (D) All three genes upregulated in male DCM patients (n = 90) relative to male CPC patients (n = 28) carried immune annotations; two (*IFI44L* and *RSAD2*) were specifically annotated with the GO term "immune response." (E) In female patients, 32 genes were DE between DCM (n = 32) and UVF (n = 50), including 23 immune-related genes. (F) None of the three DE genes in female DCM patients (n = 32) compared to female CPC patients (n = 17) carried the GO "immune response" annotation; however, *SEMA5A* (labeled) carries other immune-related annotations.

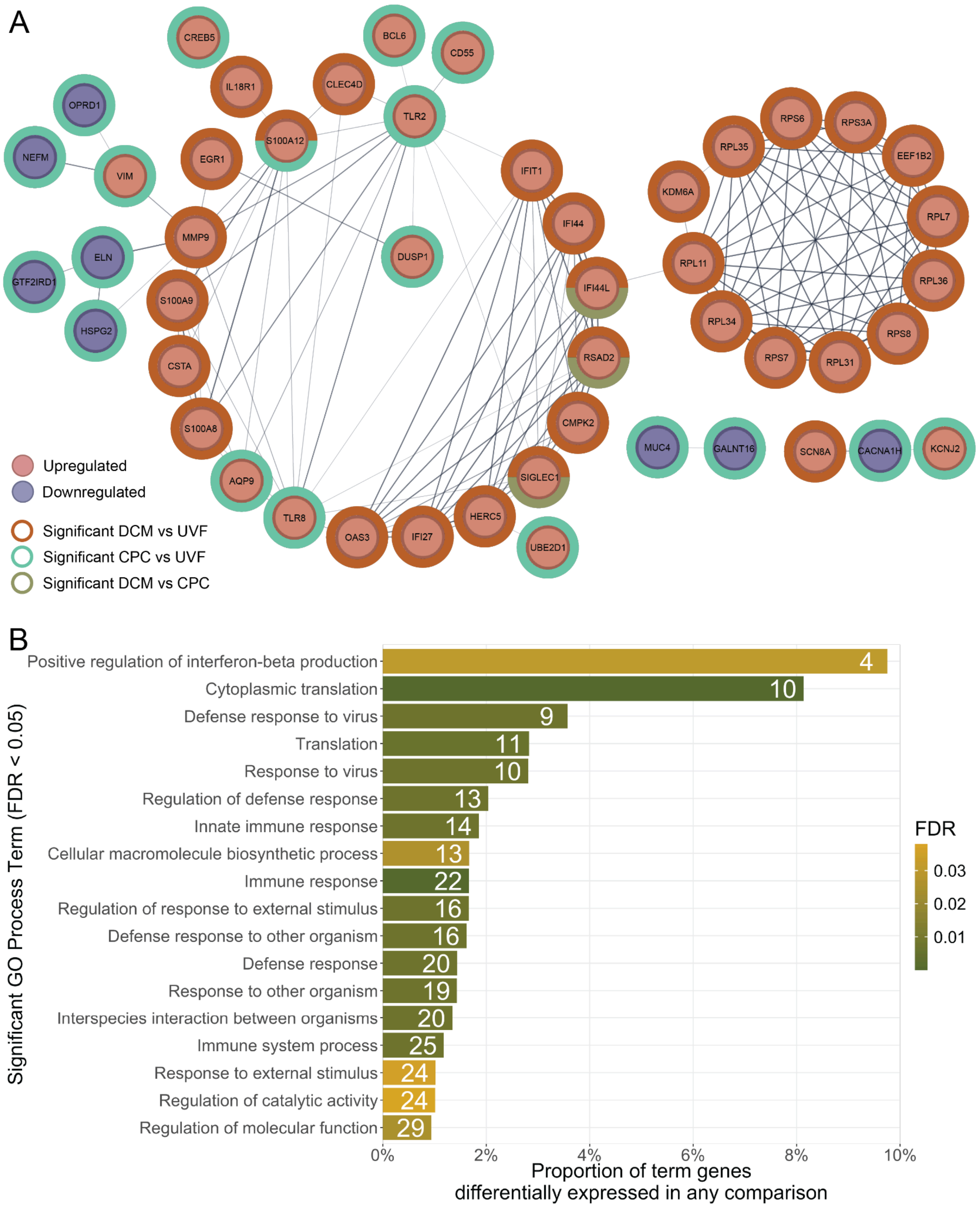


**Figure S7: Differentially expressed genes across all comparisons are enriched for immune processes.**

(A) Protein-protein interaction (PPI) network constructed from the 86 DE genes whose protein products were present in the STRING database. These genes were DE in the full cohort, in male patients only, or in female patients only. Colored rings indicate which comparisons each gene was significantly DE in (DCM vs. UVF, CPC vs. UVF, or DCM vs. CPC). Node color represents direction of effect. More than half of the proteins (48/86) had at least one PPI with another DE gene in STRING; edge thickness represents the strength of evidence for each interaction. (B) The 86 protein-coding DE genes were significantly enriched for multiple STRING functional annotations, including the 18 GO Biological Process terms shown (FDR < 0.05).

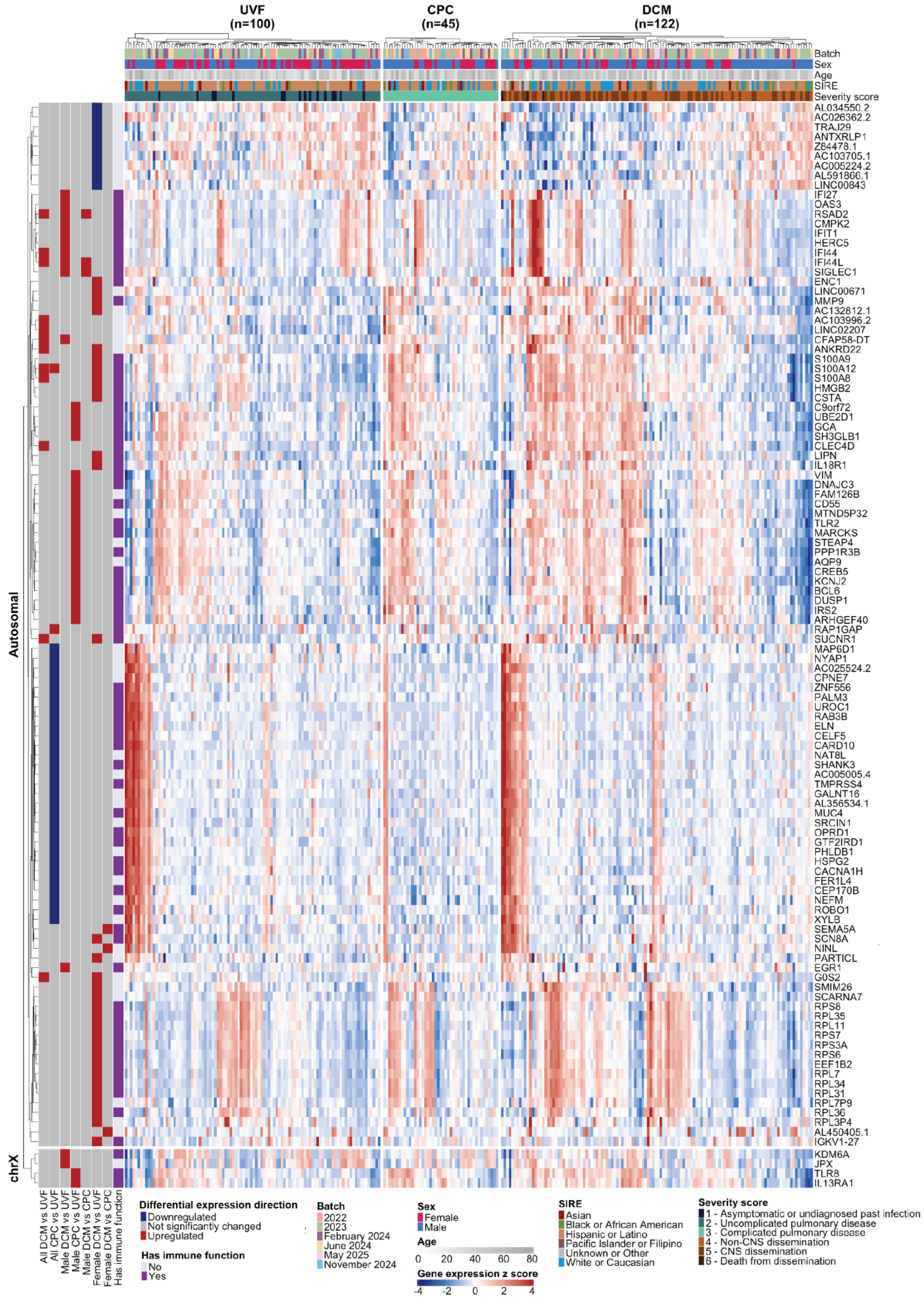
UVF
(n=100)
CPC
(n=45)
DCM
(n=122)
Batch
Sex
Age
SIRE
Severity score
AL034550.2
AC026362.2
TRAJ29
ANTXRLP1
Z84478.1
AC103705.1
AC005224.2
AL591866.1
LINC00843
IFI27
OAS3
RSAD2
CMPK2
IFIT1
HERC5
IFI44
IFI44L
SIGLEC1
ENC1
LINC00671
MMP9
AC132812.1
AC103996.2
LINC02207
CFAP58-DT
ANKRD22
S100A9
S100A12
S100A8
HMGB2
CSTA
C9orf72
UBE2D1
GCA
SH3GLB1
CLEC4D
LIPN
IL18R1
VIM
DNAJC3
FAM126B
CD55
MTND5P32
TLR2
MARCKS
STEAP4
PPP1R3B
AQP9
CREB5
KCNJ2
BCL6
DUSP1
IRS2
ARHGEF40
RAP1GAP
SUCNR1
MAP6D1
NYAP1
AC025524.2
CPNE7
ZNF556
PALM3
UROC1
RAB3B
ELN
CELF5
CARD10
NAT8L
SHANK3
AC005005.4
TMPRSS4
GALNT16
AL356534.1
MUC4
SRCIN1
OPRD1
GTF2IRD1
PHLDB1
HSPG2
CACNA1H
FER1L4
CEP170B
NEFM
ROBO1
XYLB
SEMA5A
SCN8A
NINL
PARTICL
EGR1
G0S2
SMIM26
SCARNA7
RPS8
RPL35
RPL11
RPS7
RPS3A
RPS6
EEF1B2
RPL7
RPL34
RPL31
RPL7P9
RPL36
RPL3P4
AL450405.1
IGKV1-27
KDM6A
JPX
TLR8
IL13RA1
Autosomal
chrX
All DCM vs UVF
All CPC vs UVF
Male DCM vs UVF
Male CPC vs UVF
Male DCM vs CPC
Female DCM vs UVF
Female DCM vs CPC
Has immune function
Differential expression direction
Downregulated
Not significantly changed
Upregulated
Has immune function
No
Yes
Batch
2022
2023
February 2024
June 2024
May 2025
November 2024
Sex
Female
Male
Age
0 20 40 60 80
Gene expression z score
-4 -2 0 2 4
SIRE
Asian
Black or African American
Hispanic or Latino
Pacific Islander or Filipino
Unknown or Other
White or Caucasian
Severity score
1 - Asymptomatic or undiagnosed past infection
2 - Uncomplicated pulmonary disease
3 - Complicated pulmonary disease
4 - Non-CNS dissemination
5 - CNS dissemination
6 - Death from dissemination

**Figure S8: Disease-related differential gene expression is sex-specific.**

Heatmap showing expression levels of all 112 DE genes. Rows (genes) are clustered unsupervised within autosomal and chromosome X categories; columns (samples) are clustered unsupervised within disease severity categories. The direction of effect for each pairwise comparison is indicated on the left, and sample demographic characteristics are displayed across the top.

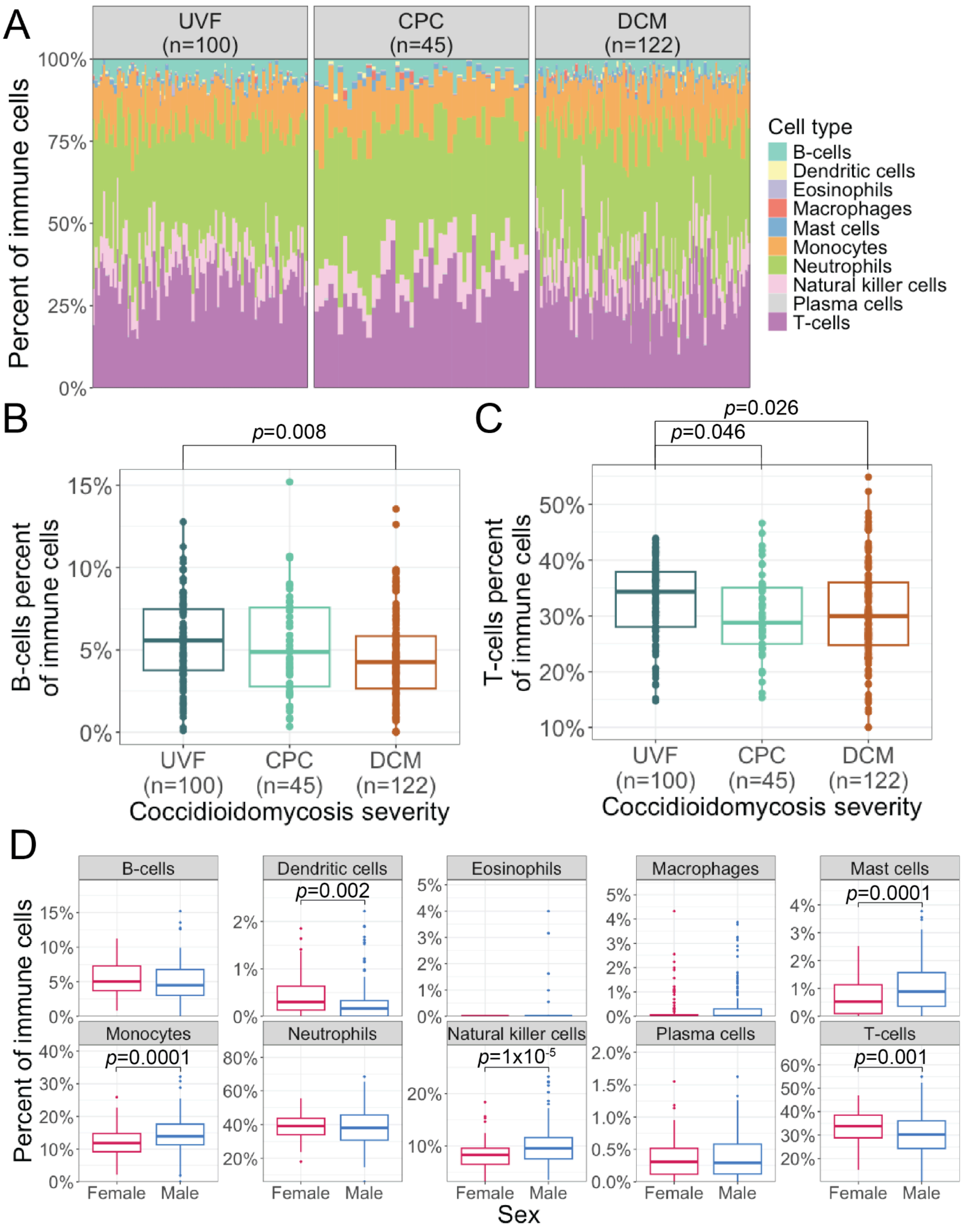


**Figure S9: Estimated immune cell type proportions by coccidioidomycosis disease severity and sex.** (A) CIBERSORTx deconvolution of blood RNA-seq data was used to estimate the proportional composition of immune cell types for each coccidioidomycosis patient. For visualization, immune cell subtypes are grouped by major lineage (e.g., naïve and memory B cells are combined under "B cells"). (B) DCM patients had a significantly lower estimated B-cell proportion than UVF patients (mean difference: −1.0%, $p$ = 0.006, Welch two-sample $t$-test). (C) UVF patients had a significantly higher estimated T-cell proportion than both DCM patients (mean difference: +2.4%, $p$ = 0.025, Welch two-sample $t$-test) and CPC patients (mean difference: +2.9%, $p$ = 0.028). (D) Five immune cell types differed significantly between male and female patients. Mast cell ($p$ = 0.00014, +0.4%), monocyte ($p$ = 0.00011, +2.4%), and natural killer cell ($p = 1.4\times10^{-5}$, +1.7%) proportions were significantly higher in males. Dendritic cell ($p$ = 0.0022, −0.2%) and T-cell ($p$ = 0.0015, −3.1%) proportions were significantly lower in males. All comparisons by Welch two-sample $t$-test.

.

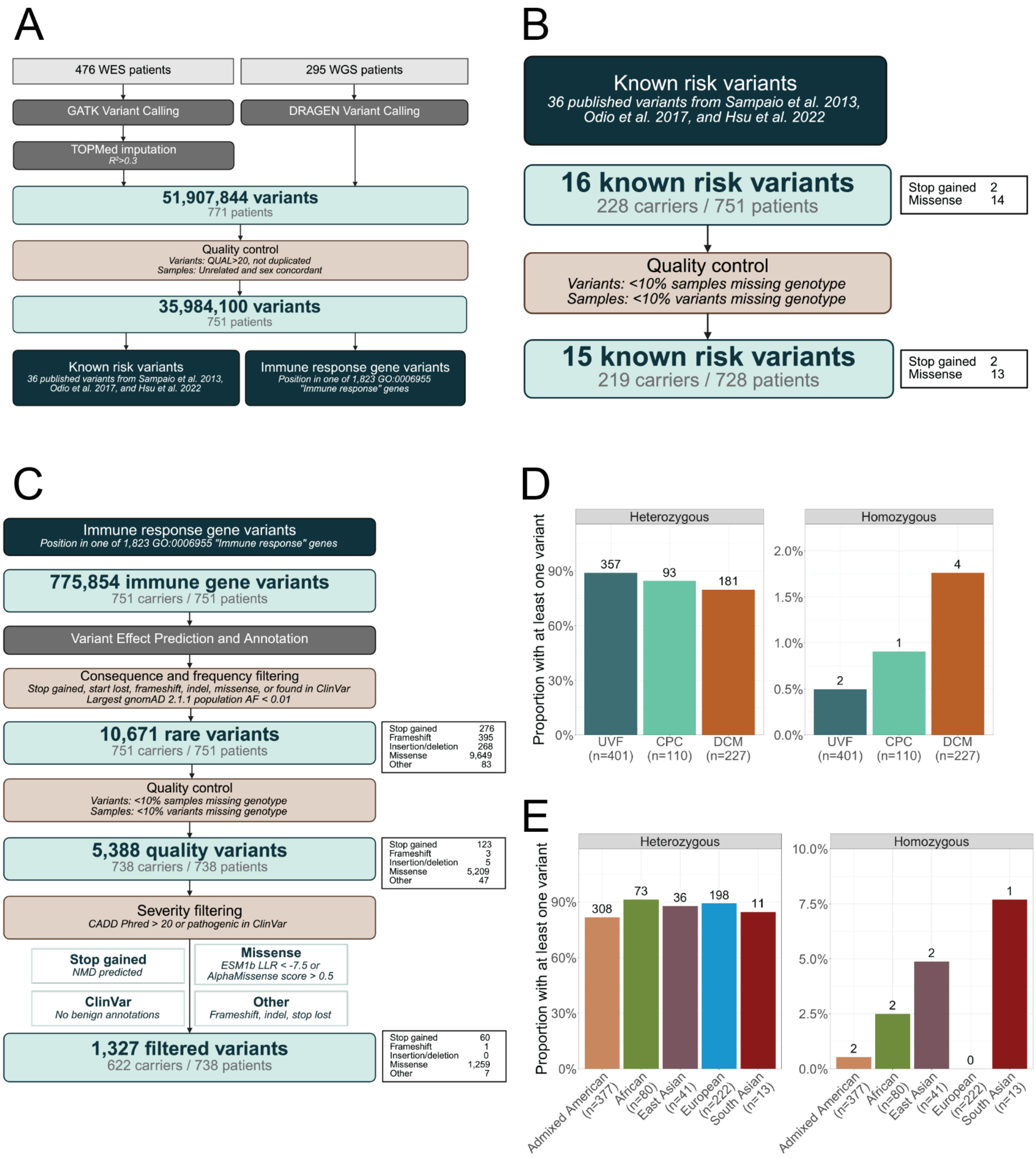


**Figure S10: Rare variant filtering workflow and overview.**

(A) Schematic of the analytical workflow used to construct the combined sequencing dataset from whole-exome (WES) and whole-genome (WGS) sequencing. WES variants were imputed and merged with WGS variants. After an initial quality control step, 751 unrelated patients whose genetic sex matched their recorded sex were retained. Two parallel rare-variant analyses were then performed. (B) We evaluated genotypes for all 36 variants previously reported in published genetic studies of coccidioidomycosis; 15 variants had carriers among the 728 patients who passed quality control for this analysis. (C) For a broader survey of rare coding variation, we focused on genes annotated with the GO term "immune response." After stringent filtering for variant type and predicted severity, 1,327 variants of interest were identified. (D) The proportion of patients carrying at least one heterozygous variant among the 1,327 filtered variants did not differ across disease severity groups. However, DCM patients had a higher proportion of individuals carrying at least one homozygous variant. (E) While patients with European genetic ancestry had similar rates of heterozygous carrier status, no patients with European genetic ancestry carried a homozygous variant.

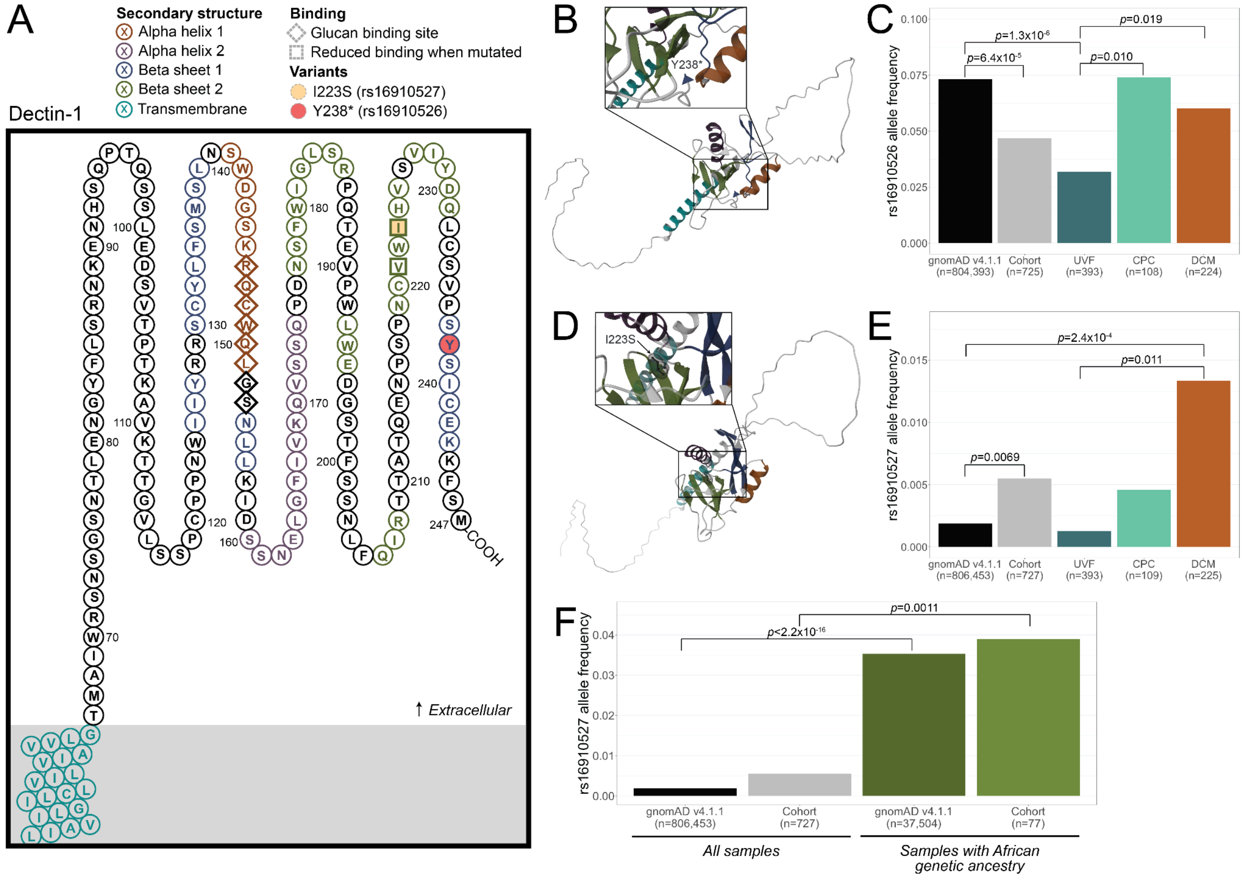


**Figure S11: *CLEC7A* variants are associated with severe coccidioidomycosis. (A)** This Protter diagram represents the extracellular amino acid sequence of the protein encoded by *CLEC7A*, Dectin-1. Two variants of interest found in our cohort are marked in yellow (p.I223S) and red (p.Y238*). Amino acids found by Brown et al. 2007 to make up the β-glucan binding site are marked with diamonds. The two amino acids that resulted in reduced β-glucan binding when mutated by Adachi et al. 2004 are marked with squares. The colors of each amino acid represent the secondary structure of the wild-type protein. **(B)** AlphaFold predicted that p.Y238* (rs16910526, labeled in inset) would disrupt a beta-pleated sheet (blue) in addition to producing a truncated protein. **(C)** While p.Y238* was significantly less common in our cohort overall than in gnomAD v4.1.1 ($p$=8.4x10$^{-5}$, Fisher's Exact Test for Count Data), the variant was significantly more common in patients with CPC ($p$=0.010) and DCM ($p$=0.019) than those with UVF. (D) Missense variant p.I223S (rs16910527, black stick diagram labeled in inset) is predicted to alter the secondary structure of an alpha helix (purple) and a beta-pleated sheet (green). It is also known to reduce β-glucan binding when mutated. (E) p.1223S is significantly more common in our cohort than in gnomAD v4.1.1 ($p$=0.0069) and in those with DCM than those with UVF ($p$=0.011). (F) The variant is also significantly more common in those with African genetic ancestry in gnomAD v4.1.1 ($p$<2.2x10$^{-16}$) and in our cohort ($p$=0.0011).

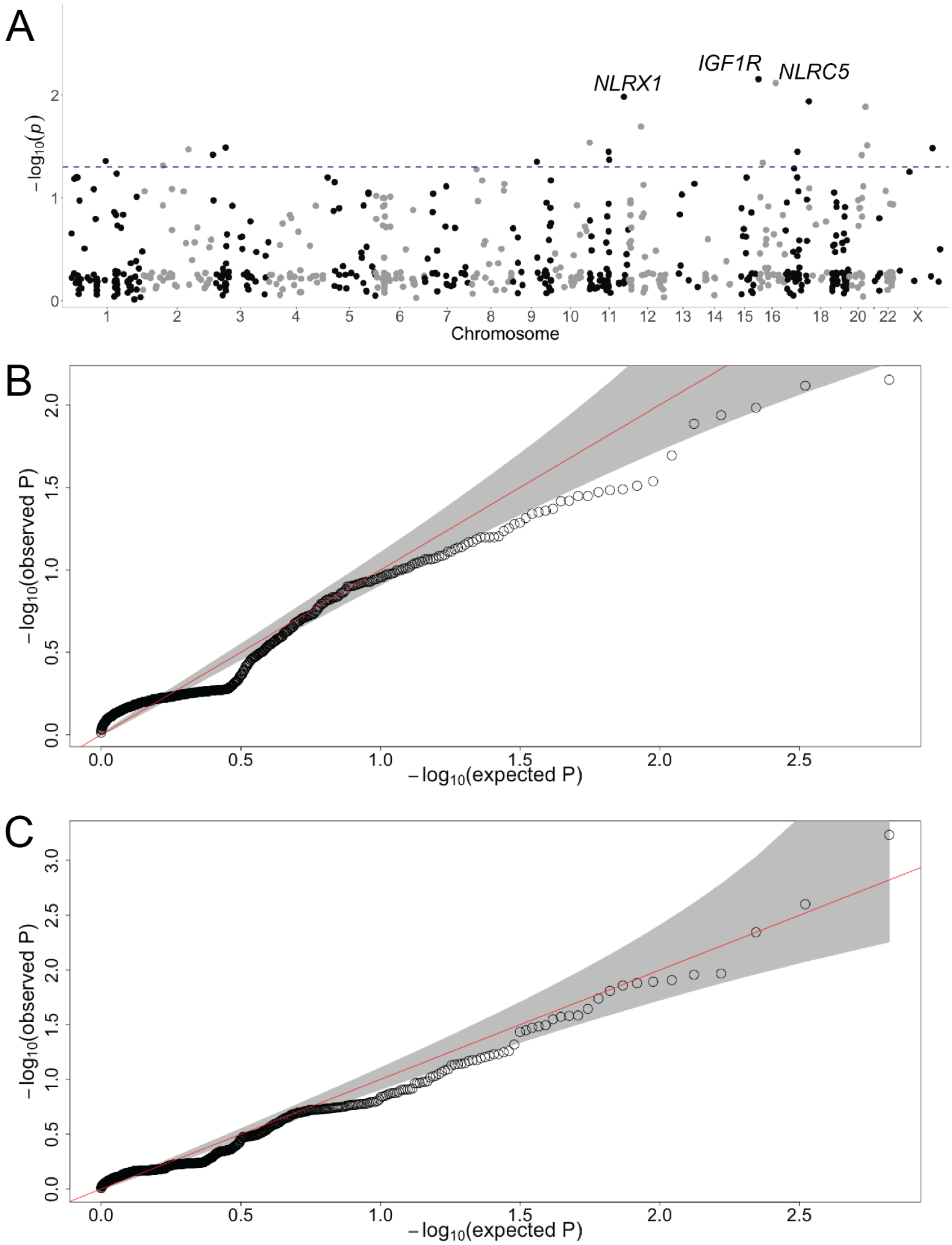


**Figure S12: Comparison of SKAT-O models shows that correcting for principal components results in *p*-value deflation.**

(A) Manhattan plot showing the -log *p*-values per gene generated by a SKAT-O model correcting for the first 10 PCs in addition to age and sex. The blue dotted line represents the nominally significant threshold ($p<0.05$). No genes reached the significance threshold after correcting for family-wise error rate (FWER, $p<5.5\times10^{-4}$). While *NLRX1* is

no longer significant after FWER correction, it is among the top 3 most significant genes.
(B) Quantile-quantile (Q-Q) plot of the expected *p*-values (x-axis) and observed *p*-values (y-axis) of a SKAT-O analysis using a model correcting for the first 10 principal components (PCs), age, and sex.
(C) Q-Q plot of SKAT-O model correcting only for age and sex.

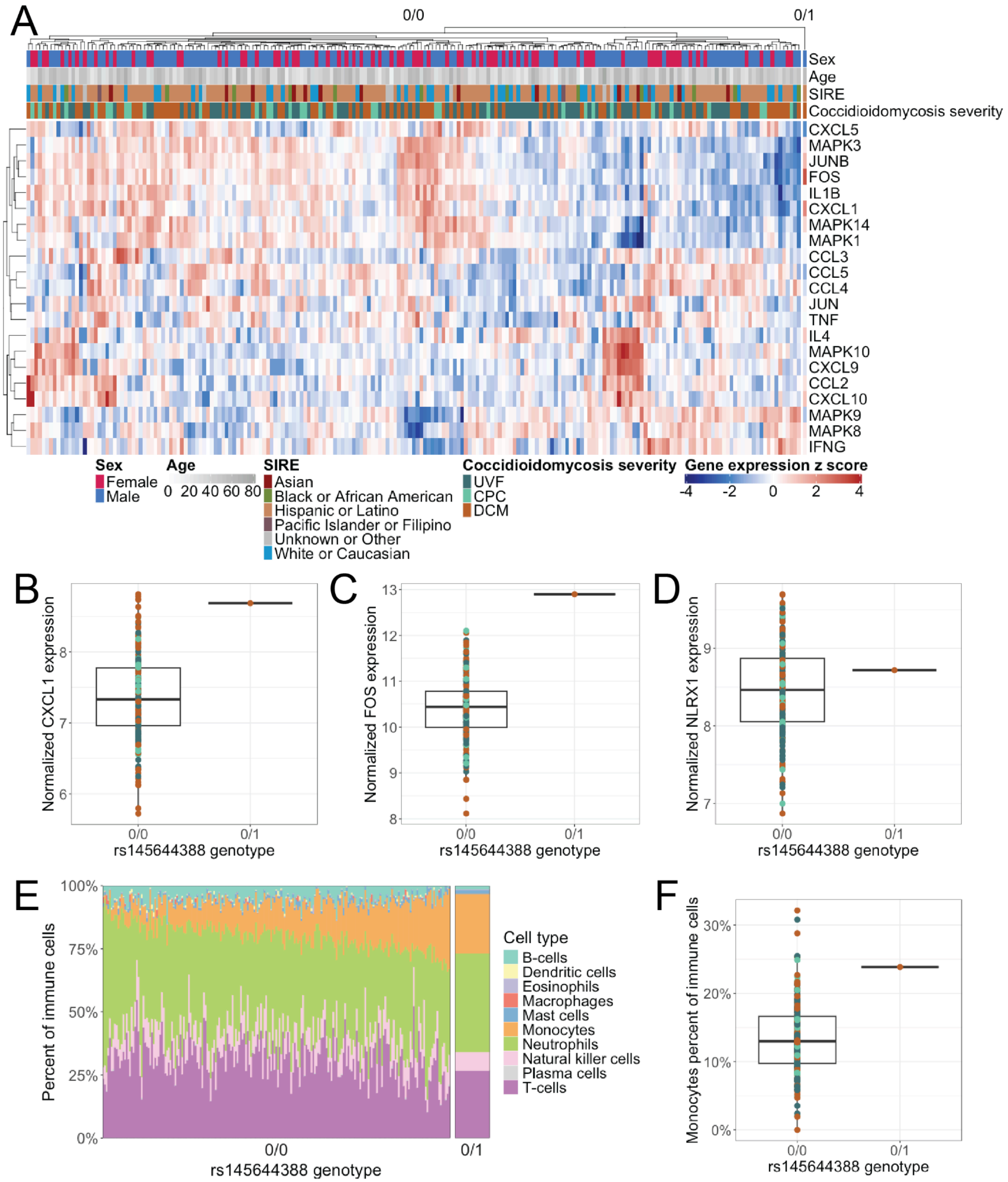


**Figure S13: Exploratory transcriptomic analysis of the *NLRX1* p.R252W (rs145644388) carrier.** (A) This heatmap represents the gene expression of cytokine, chemokine, and other immune-related genes that were differentially expressed in *NLRX1* mouse knock-out (KO) models. Only one heterozygous carrier of the variant had RNA-seq (far right column). (B) The patient with p.R252W had one of the highest expression levels of chemokine *CXCL1*. (C) The carrier also had the highest expression of *FOS*. (D) *NLRX1* expression levels are not significantly different from those without the variant, consistent with the heterozygous nature of the mutation and the low proportion of macrophages and dendritic cells in whole blood. (E) CIBERSORTx-estimated immune cell type proportions for the carrier (right) were largely similar to those of all other patients (left); most cell-type differences reported in *Nlrx1* KO mouse models were not recapitulated in this individual. (F) The carrier did have a higher than average predicted percentage of monocytes (23.9% of immune cells), although this did not reach statistical significance ($p$=0.068, linear regression controlling for age, sex, and SIRE).

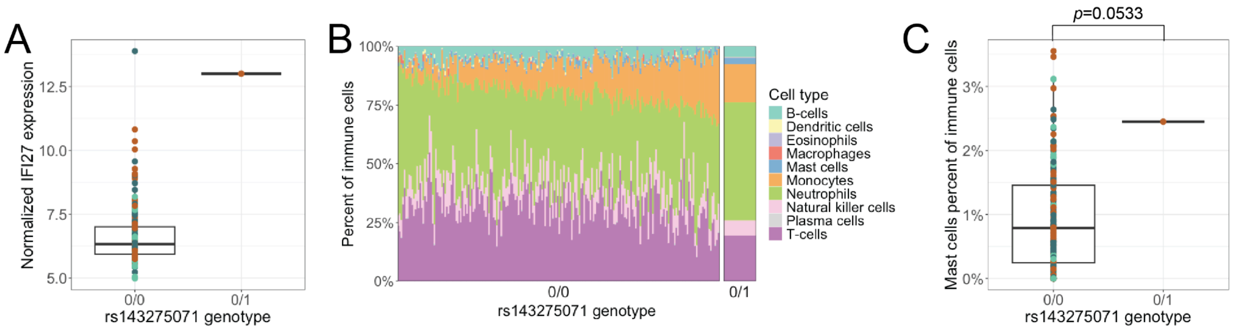


**Figure S14: *IFI27* p.Ser63Leu (rs143275071) is associated with elevated *IFI27* expression.**

(A) One patient carrying a heterozygous *IFI27* p.Ser63Leu variant had the second-highest *IFI27* expression level in the cohort, which was significantly elevated compared to non-carriers ($p$ = 1.58×10$^{-8}$, linear regression controlling for age, sex, and SIRE). (B) CIBERSORTx-estimated immune cell type proportions for the carrier (right) compared to all other patients (left) suggest a visually elevated mast cell fraction (light blue). (C) The carrier's estimated mast cell proportion (2.4% of immune cells) was higher than that of most other patients, though this difference did not reach statistical significance ($p$ = 0.067, linear regression controlling for age, sex, and SIRE).
